# Transformer-based deep learning model for the diagnosis of suspected lung cancer in primary care based on electronic health record data

**DOI:** 10.1101/2024.07.02.24309824

**Authors:** Lan Wang, Yonghua Yin, Ben Glampson, Robert Peach, Mauricio Barahona, Brendan C Delaney, Erik K Mayer

**Author notes:** Corresponding author Prof Brendan C Delaney Chair in Medical Informatics and Decision Making Imperial College IX 5th floor iHub 84 Wood Ln, London W12 0BZ., http://www.imperial.ac.uk/people/brendan.delaney. Joint senior authors (Brendan C Delaney, Erik K Mayer). **Funding** Cancer Research UK.

## Abstract

**Background:** Due to its late stage of diagnosis lung cancer is the commonest cause of death from cancer in the UK. Existing epidemiological risk models in clinical usage, which have Positive Predictive Values (PPV) of less than 10%, do not consider the temporal relations expressed in sequential electronic health record (EHR) data. Machine learning with deep ‘transformer’ models can learn from these temporal relationships. We aimed to build such a model for lung cancer diagnosis in primary care using EHR data.

**Methods:** In a nested case-control study within the Whole Systems Integrated Care (WSIC) dataset, lung cancer cases were identified and control cases of ‘other’ cancers or respiratory conditions. GP EHR data going back three years from the date of diagnosis less the most recent one months were semantically pre-processed by mapping from more than 30,000 terms to 450. Model building was performed using ALBERT with a Logistic Regression Classifier (LRC) head. Clustering was explored using k-means. We split the data into 70% training and 30% validation. An additional regression model alone was built on the pre-processed data as a comparator.

**Findings:** Based on 3,303,992 patients from January 1981 to December 2020 there were 11,847 lung cancer cases of whom 9,629 had died. 5,789 cases and 7,240 controls were used for training and a population of 368,906 for validation. Our model achieved an AUROC of 0·924 (95% CI 0·921– 0·927) with a PPV of 3·6% (95% CI 3·5 – 3·7) and Sensitivity of 86·6% (95% CI 85·3 – 87·8) based on the three year’s data prior to diagnosis less the immediate month before index diagnosis. The comparator regression model achieved a PPV of 3·1% (95% CI 3·0 – 3·1) and AUROC of 0·887 (95% CI 0·884 – 0·889).

**Interpretation:** Capturing temporal sequencing between cancer and non-cancer pathways to diagnosis enables much more accurate models. Future work will focus on external dataset validation and integration into GP clinical systems for evaluation.

**RESEARCH IN CONTEXT:** *Evidence before the study:* Predictive models for early detection of cancer are a priority as treatment intensity and cancer outcomes and survival are strongly linked to cancer stage at diagnosis. We searched PubMed and Embase for research on lung cancer prediction, using the search terms “lung cancer”, “diagnos$”, and “prediction model” between Jan 1, 2000 and Dec 31, 2023, to look into the contemporary research on prediction models for lung cancer. The QCancer Lung model has been recommended for prediction of lung cancer in primary care. However, classic regression models do not consider the rich relationships and dependencies in the electronic health record (EHR) data, such as cough followed by pneumonia rather than just cough in isolation. Since 2018, with advances in the natural language processing (NLP) domain, transformer-based models have been applied on large amounts of EHR data for clinical predictive modelling. We searched Google Scholar and PubMed for studies using transformer-based models on EHR data. We used the terms (“transformer” OR “bert” OR “pretrain” OR “prediction” OR “predictive modelling” OR “contextualised”) AND (“ehr” OR “health records” OR “healthcare” OR “clinical records” OR “cancer” OR “disease”) in free text, published from Jan 2019 to Dec 2023. We found these studies were limited to diagnosis and medication concepts/codes in patients’ records in secondary care, omitting symptom, test, procedure, and referral codes. The early detection of lung cancer requires the improvement in the prediction performance of deep learning models. We updated the literature review when writing this paper (Apr 2024) to include the latest published studies.

*Added value of this study:* We pretrained a transformer-based deep learning model, MedAlbert, for learning deep patient pathway representations from coded EHR data in primary care. This ‘Pathway to Diagnosis’ for each patient is defined to contain the most possible elaboration of the coded medical records appearing over three years before diagnosis. To our knowledge, we are the first to build models on such detailed clinical records in primary care without data aggregation. Developed and validated based on the pretrained MedAlbert, the prediction model, MedAlber+LRC, shows improved prediction performance for diagnosis of suspected lung cancer as well as one- and two-year lung cancer early detection compared with a classic machine learning model (a single Logistic Regression Model), MedAlbert+LRC performed better in terms of sensitivity, specificity, PPV and AUROC. The explainability of the model discovered a series of symptoms, comorbidities and procedures associated with lung cancer diagnosis and identified six groups of patients related to COPD, diabetes, other cancers, etc. The prediction model we developed could be applied to the UK primary care population for early diagnosis of lung cancer.

*Implications of all available evidence:* In order to progress beyond simple ‘red flag’ driven referral guidance and to develop more accurate prediction models for early diagnosis of lung cancer, it is necessary to use more sophisticated machine learning methods. Additionally, the framework we designed for deriving, modelling, and analysing the patient pathways could be used for the prediction of other cancers or diseases. The improvement in early diagnosis of lung cancer could contribute to better cancer outcomes and survival rates. Deep learning for diagnosis could provide more efficient care delivery and more accurate decisions faster, reducing costs and suffering across societies in the UK and worldwide.

## Introduction

Each year more than 45,000 UK patients are diagnosed with lung cancer,^1^ with only one third of patients diagnosed at early stage (I – II),^2^ contributing to an age standardised five-year cancer survival of only 21%.^3^ Only 4% of patients present in primary care with ‘red flag’ symptoms such as haemoptysis, most presenting with less specific features including cough or weight loss and more than a third presenting three or more times before referral.^4–6^ Robust evidence on the predictive value of combinations of symptoms and signs is limited, and based on models that do not consider the temporal evolution of codes in the electronic health record (EHR).^7,8^ Given the large volume and high dimensionality of data becoming available via integrated care systems in the UK, it is possible that Deep Learning and Natural Language Processing (NLP) approaches to EHR data analysis may provide more predictive models for early cancer diagnosis.^9^ A recent machine learning (ML) study using EHR data from 9 million patients was able to predict pancreatic cancer diagnosis within 36 months with reasonable accuracy via changes in patterns of clinical codes from the EHR. ^10^ Predictive models for lung cancer either identify prevalent risk factors such as age and smoking history for identifying at risk populations for screening,^11,12^ or add incident symptoms for use in the diagnosis of suspected cancer for referral.^8^ A systematic review of the latter found 13 studies, with haemoptysis found to have the greatest diagnostic value, diagnostic odds ratio (DOR) 6·39 (3·32 – 12·28), followed by dyspnoea 2·73 (1·54 – 4·85) then cough 2·64 (1·24 – 5·64) and chest pain 2.02 (0·88 – 4·60). Other studies have identified weight loss, anaemia, and thrombocythemia as potential predictors.^13,14^ Of the population studies, age, sex, sociodemographic factors, smoking history (recorded in a variety of ways), family history, occupational exposure, COPD, alcohol and body mass index have been included in models.

EHR data is not only noisy and heterogenous but also sparse, since typically only one or two codes are chosen by the clinician and the text note is not usually available on account of the risks to privacy. ^15,16^ The sequential relationships among presenting symptoms, referrals and tests will differ between patients with lung cancer and without and this can be used to derive an ML model. Treating sequences of structured medical data (codes) as an NLP problem unlocks more powerful ML tools. Learning context requires the model to have a long-term memory. Recurrent Neural Networks (RNNs), Long Short-Term Memory (LSTM) and gated recurrent neural networks are designed for sequence modelling and have been used to model the temporal evolution of EHR data for disease-prediction problems.^17^ However, RNNs are incapable of handling long-term dependencies because they are biased by most recent inputs in a sequence, and the sequential nature of RNN and LSTM models makes them computationally inefficient for handling large data sets. BERT (Bidirectional Encoder Representations from Transformers) uses a multi-layer bidirectional transformer encoder which enables pre-trained deep bidirectional representations by jointly conditioning on both left and right context in all layers.^18^ Furthermore, the multi-layer Transformer based architecture with a multi-head self-attention mechanism not only enables parallel computation which facilitates long-range dependency learning, but also gives the model greater power to encode a broad range of relationships and nuances for each token, for example the order of a sequence by embedding the specific position of each token in the sequence. The above properties make the model very powerful in encoding linguistic regularities and patterns and capturing precise syntactic and semantic word relationships. BERT-based models have been applied on large amounts of EHR data for clinical predictive modelling but limited to diagnosis and medication codes in patients’ records, omitting symptom, test, and referral codes. ^10,19,20^ We propose a novel framework for deriving, modelling, and analysing the entire coded patient pathways leading to the point of diagnosis with lung cancer. This allows us to discover lung cancer progression patterns and clinical investigation patterns and results in a state-of-the-art prediction model for diagnosis of lung cancer.

## Methods

### Data

We used the primary care dataset of Whole Systems Integrated Care (WSIC) Northwest London EHR data, consisting of primary care coded EHR data of patients from 400 GP practices. The records include demographic data, date of birth, gender and ethnicity, as well as episode data, patient visits to GPs, clinics, and hospitals, including medication history, diagnosis, symptoms and signs, tests and procedures coded using Read CT v2 (as shown in 1A). Owing to the lack of a standard to define whether an item is missing or not, imputation of missing data was not possible. The WSIC data was first partitioned at random into 70% for training and 30% for validation. To create a balanced dataset to train the model, we created a nested case-control study within the training set. The control subjects were over-selected to include both cancers other than lung, and respiratory conditions, to ensure the model was built to maximise its ability to detect differences between patient pathways in these conditions.

### Defining patient level pathways to diagnosis

Lung cancer patients were identified from the data using codes in Table S1. We defined a patient pathway to diagnosis as the sequence of medical codes appearing over three years before diagnosis with a temporal order ( 1D). For a patient diagnosed with lung cancer, the endpoint of the pathway was the date of the first lung cancer diagnostic code in the EHR. Lung cancer diagnoses took precedence if the patient was diagnosed with multiple cancers. We derived a pathway for each patient by working backward from the diagnosis date, listing all medical events in the EHR over three years prior to the diagnosis in an order of time, as shown in Figure 1B. To construct the control group, patients were selected based on sets of codes for ‘cancers not lung’ with the date of the first cancer diagnostic code and ‘other diagnostic codes’ using the most recent date as the endpoint of the pathway.

**Figure 1.**
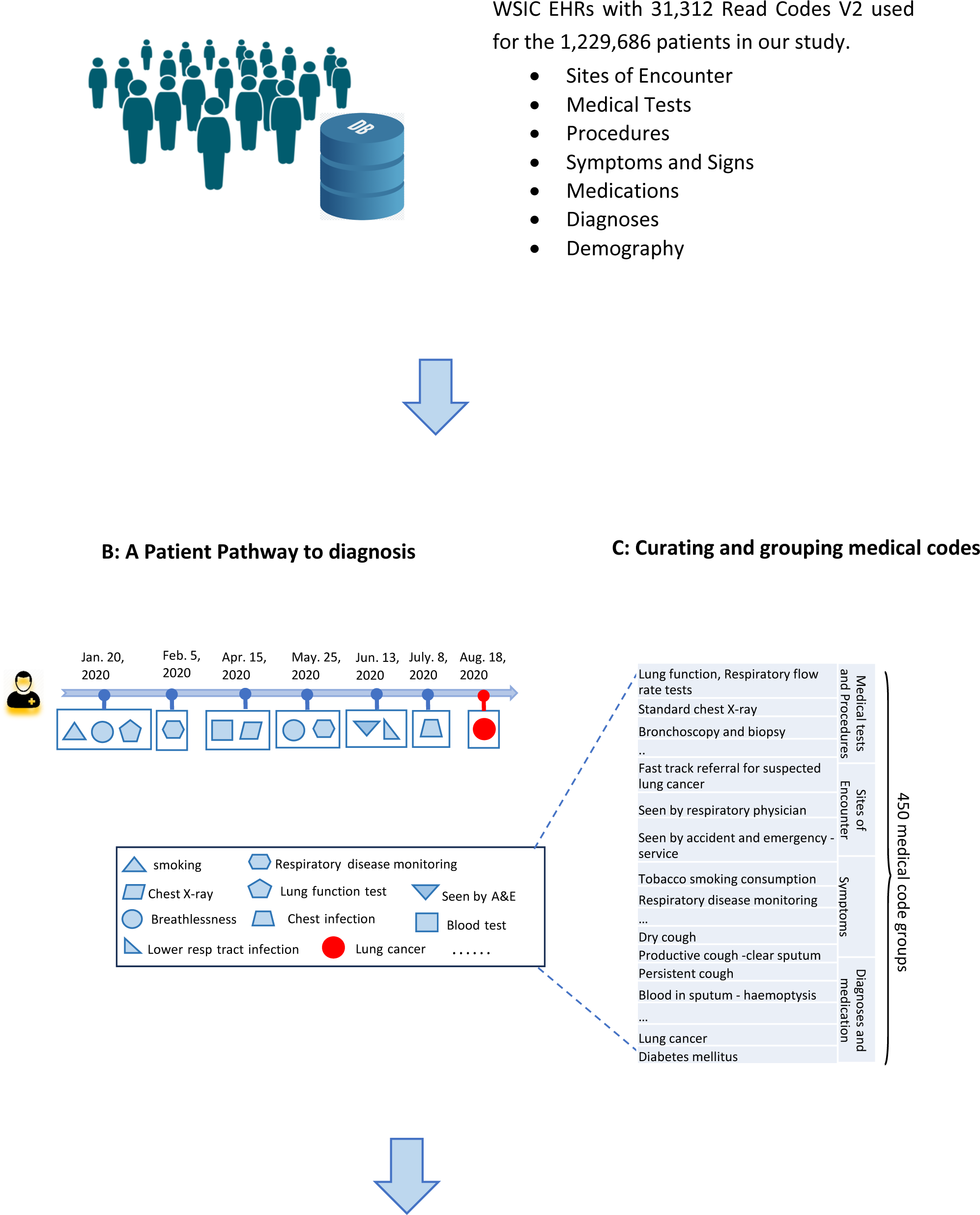
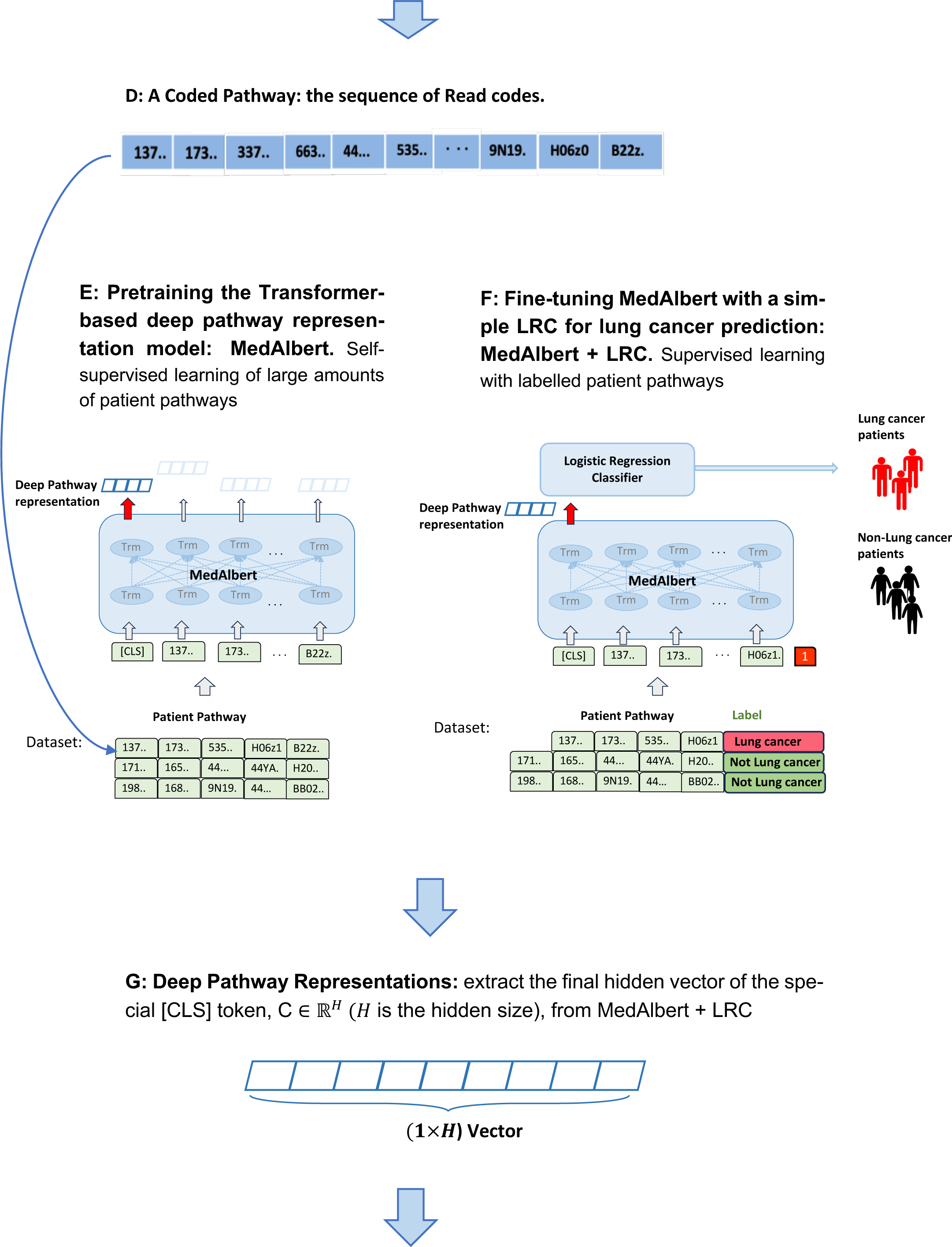
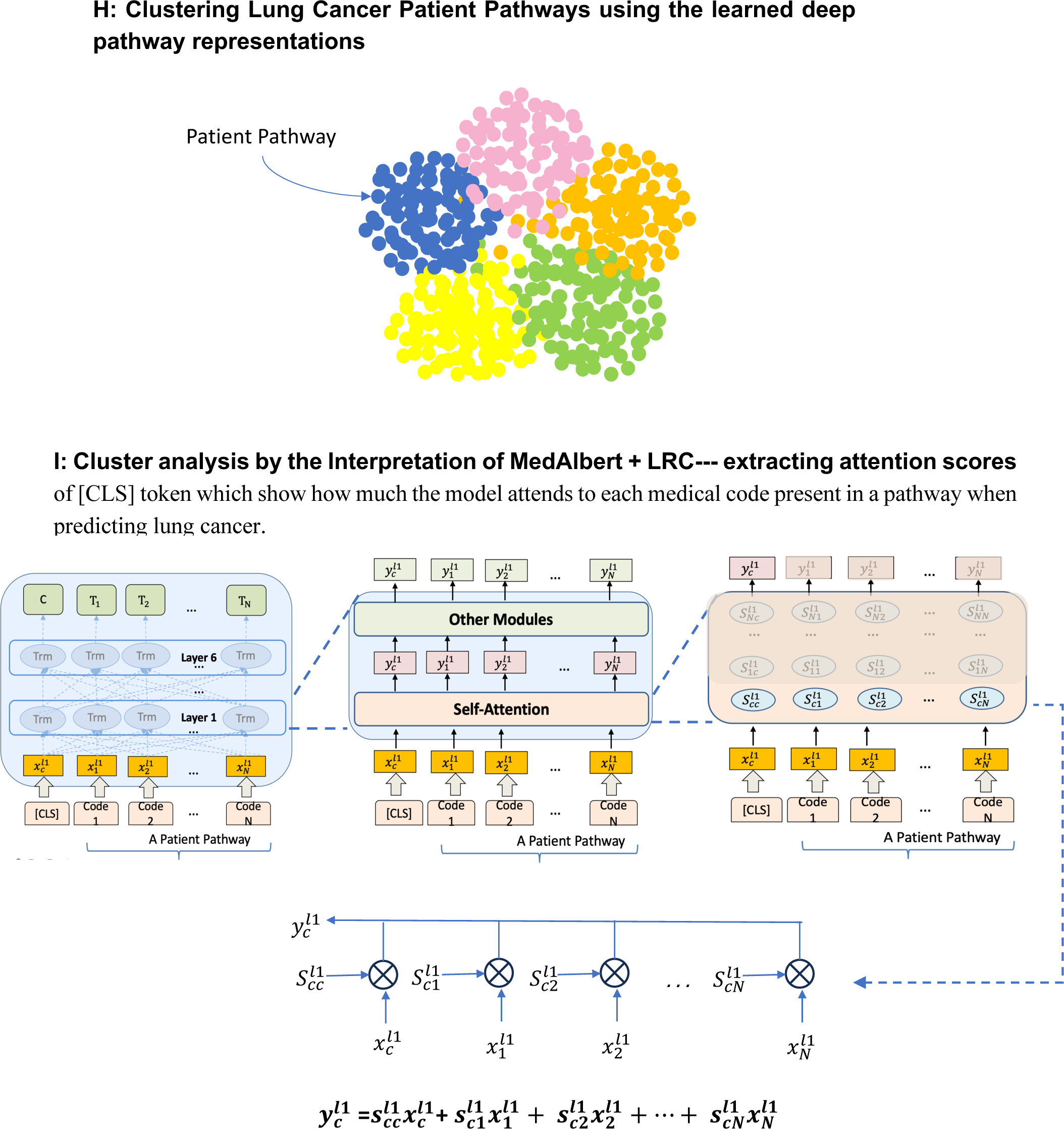
Conceptual framework of our proposed method for the diagnosis of suspected lung cancer.

**Figure 2.**
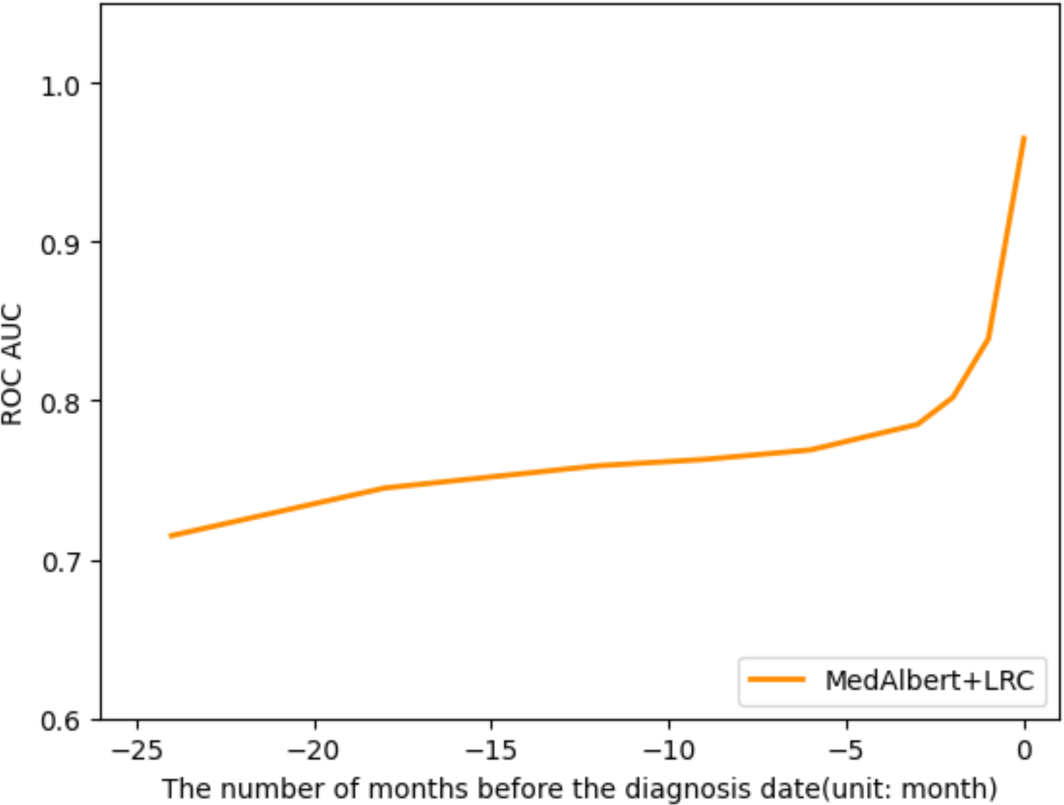
The ROC AUCs of MedAlbert+LRC tested using the pathways excluding varied time periods of medical codes presented before diagnosis date.

**Figure 3.**
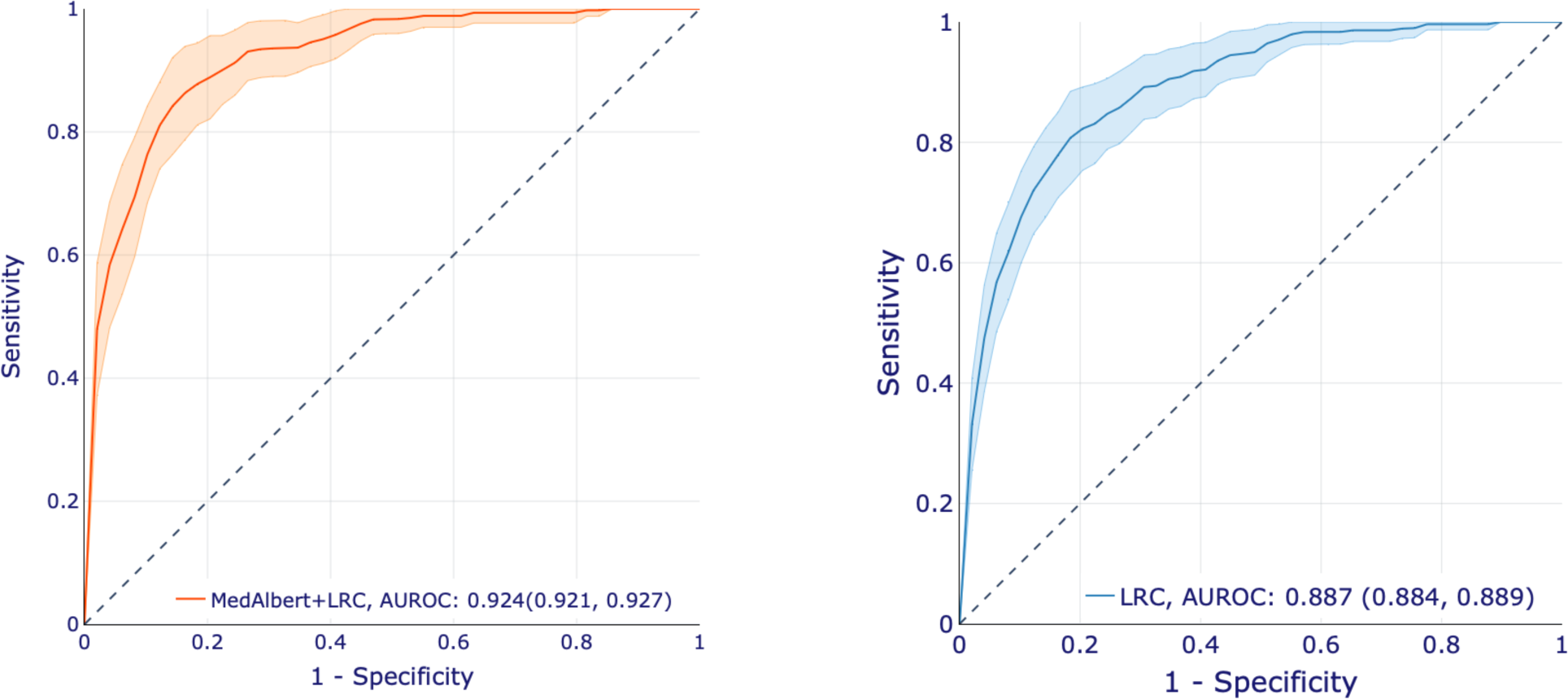
ROC curve of MedAlbert+ LRC (left) and a single LR classifier (right) applied on three-year patient pathways before diagnosis. Models are trained on three-year pathways excluding the most recent one-month codes before diagnosis.

### Curating and grouping medical codes

Amongst the 8,416 patients with lung cancer and 1,221,270 patients with other conditions there were 31,312 unique Read codes collected from their pathways. Efficient modelling requires dimension reduction in the code space. This was accomplished by clinically guided mapping up of codes to higher terms in the Read hierarchy and removal of purely administrative terms, resulting in 450 Read code groups (as shown in Figure 1C). See supplementary Material.

### Machine learning approach

#### A deep contextualised pathway representation model: MedAlbert

We designed a deep pathway representation model, MedAlbert, based on a state-of-the-art NLP model, A Lite BERT (ALBERT), ^21^ with fewer parameters and lower memory consumption than BERT. Our model, MedAlbert, uses a six-layer Transformer with twelve attention heads to learn the representations for each medical code at each layer by integrating long range (left and right) contextual information in a pathway (Figure 1E). Multi-head attention allows the attention module to repeat its computation multiple times. Therefore, there are at least 6×12 Attention calculations. Through this repeated composition of medical code embeddings, our model can learn different aspects of representations which capture a wide variety of relationships and dependencies between medical codes and form very rich representations. The final hidden state corresponding to the first input token is used as the aggregate pathway representation for Lung cancer prediction. We subsequently fine-tuned the model with a Logistic Regression Classifier to create a model for lung cancer prediction.

#### Input/Output Representations

The input and its embedding from our model are distinct from previous published work using NLP approaches to diagnostic prediction (). The input is the sequence of *N* medical codes present in a three-year pathway prior to diagnosis date for each patient with the temporal order, starting with a special token ([CLS]). Unlike previous approaches utilising the hierarchical nature of structured EHR data, being a sequence of visits over time for each patient and each visit containing multiple medical codes,^19,20^ we flattened the structured EHR into a single dimensional sequence in order to retain to the largest extent the causal relationships between medical codes recorded during a patient’s medical history. The input embedding is constructed by combining the corresponding token and position embeddings. Position embeddings encode the specific position of each medical code in the input pathway to capture the sequential relationships among codes.

#### Pre-training MedAlbert

We pre-trained MedAlbert using masked language model (MLM) based on the original implementation described in the BERT paper to enable the representations to fuse the left and the right context and, as a result, pre-train a deep bidirectional representation model. ^18^ As suggested in the BERT paper,^18^ we masked 12% of the medical codes in each input pathway at random and replaced them by [MASK], 1·5% of codes are replaced with random codes. Then, the final hidden vectors corresponding to the masked codes are used to predict the original codes. We used three-year pathways of the patients in the training dataset to pre-train MedAlbert with each pathway ending by the diagnosis code (Figure 1E). We used the default hyperparameter setting of ALBERT.

#### Lung Cancer Predictive Model

To predict the probability that an individual patient might develop lung cancer given his/her historical pathway data, we formalised lung cancer prediction as a binary classification task. We developed a deep predictive model by fine-tuning the pretrained MedAlbert with a logistic regression classifier. In addition, as a comparator, we also created a Logistic Regression (LR) classifier using the medical codes directly as input features (instead of learnt sequence representations) (see Supplementary material).

#### Deep Predictive Model: MedAlbert+LRC

The deep predictive model for lung cancer diagnosis was created by layering a Logistic Regression Classifier (LRC) on top of the output of the pre-trained Deep Contextualized Pathway Representation Model for the special [CLS] token (Figure 1F). All parameters were then jointly fine-tuned for the lung cancer prediction task. The final layer representation of [CLS], *C* ∈ ℝ^H^, is used as the aggregate pathway representation and passed to a LRC for lung cancer prediction (binary 0 or 1). ^23^ The additional parameters introduced for fine-tuning are classifier layer weights *W* ∈ ℝ^H^. We computed a Binary Cross Entropy loss with BCELoss(Sigmoid(CW^T^). The parameters of MedAlbert+LRC were fine-tuned using labelled three-year pathways (from the training set excluding the lung cancer diagnosis codes as shown in Table S1) with each pathway labelled with a respective cancer diagnosis, “1” for lung cancer diagnosis and “0” for non-lung cancer (Figure 1F). We trained with batch size of 8 pathways and 4 epochs, and other hyperparameters are the same as in pre-training. We used 600 pathways randomly selected from the training dataset as the evaluation dataset. The above optimal hyperparameter values were selected on the Eval set. Additionally, as fine-tuning was sometimes unstable, we ran several random restarts with the same pre-trained checkpoint but different training data shuffling and classifier layer initialization and selected the model that performed best on the evaluation set. ^18^ To examine the sensitivity of the models we developed to the length of the pathway, we predicted the diagnosis of lung cancer using three different types of pathways: (1) three-year pathways; (2) the first two years of the three-year pathways; (3) the first one year of the three-year pathways. (see Supplementary material) We compared the results with the classic ML model, Logistic Regression classifier (LR).

#### Evaluation Metrics

The validation metrics we used are precision, recall, and area under the ROC curve (AUROC). “Precision” shows the proportion of the patients with predicted lung cancer that are correctly predicted as lung cancer by the model as proportion of predicted +ve cases, in medical diagnostic studies this is termed, Positive Predictive Value (TP/(TP+FP). “Recall” shows the proportion of actual lung cancer patients that are correctly predicted as lung cancer as a proportion of lung cancer cases, in other words, correct diagnoses or Sensitivity (TP/(TP+FN).

#### Exploration of the impact of bias in pathway representations

When using EHR data from primary care it is possible the diagnosis dates were recorded with some delay because of the time taken to receive an email or postal communication from a hospital clinic and miss-coding of the diagnosis date in the GP record. Although it is best practice to code this on the clinic date rather than the date received, this relies on human intervention at coding. Being aware of the cancer diagnosis may affect the coding of symptoms post-diagnosis as symptoms are more likely to be coded (as opposed to being entered as free text) when they support an existing or presumed diagnosis than when their significance is uncertain. ^16^ The effect of this potential bias would be to overestimate the performance of the model. In addition, results of definitive diagnostic investigations taken after referral from primary care may also appear in the record. Potential bias can be explored by removing data for the period immediately before the diagnosis index date. To determine the appropriate number of months of data to remove, whilst still generating the optimal model we constructed a set of test datasets with removal of none, one month, and three months of data. The selected model was chosen on the prior specified criteria of being the data cut before any steep change in model prediction.

#### Cluster Analysis of Lung Cancer Patient Pathways

We aimed to investigate clinical interpretability of the MedAlbert + LRC model by using k-means clustering to examine the outputs of the trained model (Figure 1H) and exploring the attention scores. The outputs of the trained MedAlbert + LRC included the embedding of [CLS] which we use as the representation for each pathway (Figure 1G) and the attention scores for [CLS] token which show how much the model attends to each medical code present in a pathway when predicting lung cancer. We extracted the attention scores of each medical code for [CLS] in a pathway by averaging the scores over 12 attention heads at the sixth layer and, as a result, form a 1 × *N* vector (*N* is the number of medical codes in the pathway) which shows how much one code is related to each of the other codes for lung cancer prediction (as shown in Figure 1I).

## Results

In the period from January 1981 to December 2020 there were in total 3,303,992 patients in WSIC, where in December 2020, 1,980,821 were registered, 224,681 had died, and 1,098,490 had left the area. Among all the patients, 11,847 were diagnosed with lung cancer where 9,629 died, 1,306 were still registered, and 912 had left the area. To train our deep model effectively, we required a minimum number of ten medical codes we have curated in the three years before diagnosis, leaving 8,416 lung cancer patients in which 981 patients were still registered. 1,221,270 patients consulted a GP in the time period of the study with a reason other than lung cancer. The nested case-control population consisted of 5,789 lung cancer patients (44·4%), and 7,240 controls made up of 2,932 (22·5%) patients with chronic respiratory conditions, 2,030 (15·6%) patients with other cancers, and 2,279 (17·5%) patients with a wide range of other conditions (as shown in Figure S3). We pre-trained MedAlbert with batch size of 2 pathways for 390,870 steps, which is approximately 60 epochs over the 13,029 patient pathways. The clinical characteristics of the training and validation sets are shown in Table 2. The mean age at lung cancer diagnosis was 71·5 while the mean age in the whole study population was 51·4 and in the control group was 58·5.

**Table 1.** Comparison of modelling approach of MedAlbert with BEHRT,^20^ Med-BERT,^19^ DNPR model,^10^ and Foresight. ^22^ DNPR = Danish National patient Registry.

|  | BHERT | Med-BERT | DNPR model | Foresight | MedAlbert |
| --- | --- | --- | --- | --- | --- |
| Type of input codes | Caliber code for diagnosis developed by a college in London | ICD-9 + ICD-10 code for diagnosis | ICD-8 + ICD-10 code for diagnosis | Snomed for disorder, substance, finding, and procedure | Read codes v2 for symptoms, diagnoses, medications, procedures, sites of encounter, and medical tests |
| Input embeddings | Code + visit + age embeddings | Code + visit + code serialisation embeddings | Code + Age embeddings | Code + Age embeddings | Code embeddings |
| Training sample unit | Patient's visit sequence | Patient's visit sequence | Patient's code sequence | Patient's code sequence, prepending age, sex, and ethnicity, appending [SEP] between codes of each day | Patient's code sequence |
| Vocabulary size | 301 | 82,000 | 2997 | 19,5416 | 450 |
| Average number of visits for each patient for pretraining | Not reported but >5 | 8 | 18-121 | Not reported but < 256 | 170 |
| Minimum visits per patient | 3 | 5 | 5 | 10 | 10 |

**Table 2.** Demographic and clinical characteristics of the training + evaluation, and validation cohorts for MedAlbert and MedAlbert + LRC.

|  | WSIC whole population<br>n (%) | Training cases n (%)<br>(lung cancer) | Training controls n (%)<br>(non-lung cancer) | Validation set n (%) |
| --- | --- | --- | --- | --- |
| Dataset size | 1229686 | 5789 (0·5%) | 7240 (0·6%) | 368906 (30·0%) |
| Lung cancer cases | 8416 (0·7%) | 5789 (0·5%) | NA | 2627 (0·2%) |
| Age groups |  |  |  |  |
| <25 | 137185 (11·2%) | 9 (0·2%) | 985 (13·6%) | 40994 (11·1%) |
| 25-34 | 152168 (12·4%) | 25 (0·4%) | 292 (4·0%) | 46100 (12·5%) |
| 35-44 | 212612 (17·3%) | 51 (0·9%) | 549 (7·6%) | 63825 (17·3%) |
| 45-54 | 179059 (14·6%) | 358 (6·2%) | 699 (9·7%) | 53614 (14·5%) |
| 55-64 | 178166 (14·5%) | 971 (16·8%) | 1135 (15·7%) | 53623 (14·5%) |
| 65-74 | 154934 (12·6%) | 1832 (31·7%) | 1350 (18·6%) | 46195 (12·5%) |
| 75-84 | 125505 (10·2%) | 1834 (31·7%) | 1273 (17·6%) | 37260 (10·1%) |
| 85-94 | 74763 (6·1%) | 673 (11·6%) | 857 (11·8%) | 22650 (6·1%) |
| 95-104 | 12489 (1·0%) | 32 (0·6%) | 87 (1·2%) | 3721 (1·0%) |
| >105 | 2770 (0·2%) | 3 (0·1%) | 7 (0·1%) | 853 (0·2%) |
| Gender |  |  |  |  |
| Female | 750781 (61·1%) | 2859 (49·4%) | 4087 (56·4%) | 225533 (61·1%) |
| Male | 478863 (38·9%) | 2930 (50·6%) | 3153 (43·6%) | 143536 (38·9%) |
| Ethnicity |  |  |  |  |
| White-British | 324576 (26·4%) | 2527 (43·7%) | 3378 (46·7%) | 99757 (27·0%) |
| White-Irish | 28869 (2·3%) | 454 (7·8%) | 191 (2·6%) | 9019 (2·4%) |
| White-Other | 248506 (20·2%) | 1260 (21·8%) | 1054 (14·6%) | 76782 (20·8%) |
| Indian | 181586 (14·8%) | 418 (7·2%) | 1051 (14·5%) | 55771 (15·1%) |
| Pakistani | 42059 (3·4%) | 81 (1·4%) | 201 (2·8%) | 12999 (3·5%) |
| Bangladeshi | 11208 (0·9%) | 28 (0·5%) | 43 (0·6%) | 3358 (0·9%) |
| Other Asian | 102650 (8·3%) | 199 (3·4%) | 452 (6·2%) | 31529 (8·5%) |
| Caribbean | 36431 (3·0%) | 181 (3·1%) | 131 (1·8%) | 10930 (3·0%) |
| Black African | 51143 (4·2%) | 73 (1·3%) | 144 (2·0%) | 15351 (4·2%) |
| Chinese | 10831 (0·9%) | 54 (0·9%) | 21 (0·3%) | 3247 (0·9%) |
| Other | 163113 (13·3%) | 391 (6·8%) | 564 (7·8%) | 50107 (13·6%) |
| Alcohol and Smoking |  |  |  |  |
| Drinker | 506875 (41·2%) | 2851 (49·2%) | 2517 (34·8%) | 150496 (40·8%) |
| Non-drinker | 84621 (6·9%) | 381 (6·6%) | 503 (7·0%) | 25259 (6·8%) |
| Drinking-Unknown | 613 (0·0%) | 0 (0·0%) | 9 (0·1%) | 184 (0·0%) |
| Ex-drinker | 6869 (0·6%) | 57 (1·0%) | 14 (0·2%) | 2429 (0·7%) |
| Non-smoker | 638874 (52·0%) | 1551 (26·8%) | 3475 (48·0%) | 205664 (55·7%) |
| Ex-smoker | 287932 (23·4%) | 3130 (54·1%) | 2551 (35·2%) | 84076 (22·8%) |
| Smoker | 213579 (17·4%) | 2748 (47·5%) | 1383 (19·1%) | 63806 (17·3%) |
| Cancers |  |  |  |  |
| Oral cancer | 1568 (0.1%) | 69 (1.2%) | 43 (0.6%) | 415 (0.1%) |
| Gastric-oesophageal cancer | 2279 (0.2%) | 57 (1.0%) | 70 (1.0%) | 602 (0.2%) |
| Colorectal cancer | 6865 (0.6%) | 262 (4.5%) | 183 (2.5%) | 1840 (0.5%) |
| Pancreatic cancer | 1637 (0.1%) | 34 (0.6%) | 46 (0.6%) | 420 (0.1%) |
| Skin, bone, connective tissue cancer | 2662 (0.2%) | 102 (1.8%) | 61 (0.8%) | 693 (0.2%) |
| Breast cancer | 10097 (0.8%) | 402 (6.9%) | 199 (2.7%) | 2170 (0.6%) |
| Uterine cancer | 1971 (0.2%) | 55 (1.0%) | 47 (0.7%) | 530 (0.1%) |
| Ovary cancer | 1299 (0.1%) | 21 (0.4%) | 44 (0.6%) | 350 (0.1%) |
| Cervical cancer | 506 (0.0%) | 27 (0.5%) | 11 (0.2%) | 133 (0.0%) |
| Prostate cancer | 11398 (0.9%) | 220 (3.8%) | 392 (5.4%) | 3217 (0.9%) |
| Renal cancer | 4451 (0.4%) | 162 (2.8%) | 90 (1.2%) | 1198 (0.3%) |
| Brain cancer | 627 (0.1%) | 25 (0.4%) | 24 (0.3%) | 159 (0.0%) |
| Thyroid cancer | 949 (0.1%) | 28 (0.5%) | 26 (0.4%) | 264 (0.1%) |
| Blood cancer | 7560 (0.6%) | 124 (2.1%) | 260 (3.6%) | 2818 (0.8%) |

### Deep representations of patient pathways improve lung cancer prediction

Sensitivity analysis to determine the amount of data to be removed immediately prior to the index date was conducted. The results are shown in . The performance of the model increased steadily when more medical codes before the diagnosis date are included in the pathways. After the time point of three months, and again at one month, the ROC AUCs rose. This might result from two reasons, bias, or the possibility that the symptoms and attendances of the patients become more predictive when approaching diagnosis. For a conservative approach to avoiding bias we selected to pretrain our deep pathway representation model, MedAlbert, and then fine-tune MedAlbert with a LRC, using the three-year pathways excluding the one month of data immediately preceding the index diagnosis.

Extensive sensitivity analysis was conducted as to the impact of the amount of clinical data included before the index date and the impact of trimming the immediate period before the index diagnosis. Table 3 and Table S3 –Table S10 present the comparison of Precision, Recall, F1-score, and AUROC between the two predictive models (MedAlbert+LRC and LR) trained and tested on the nine combinations of years of data and number of months excluded before the end date. The ROC curves are shown in Figure 4 and Figure S4 – Figure S6. Our chosen model was trained by the three-year data with one month removed. The selected MedAlbert+LRC always outperforms the single LR by a substantial margin, obtaining a 1% – 6% absolute improvement in Precision, Recall, F1 score, and AUROC. In particular, the selected model shows quite good performance in one- and two-year early diagnosis of lung cancer, achieving AUROC of 86·3% and 83·3% respectively.

**Figure 4.**
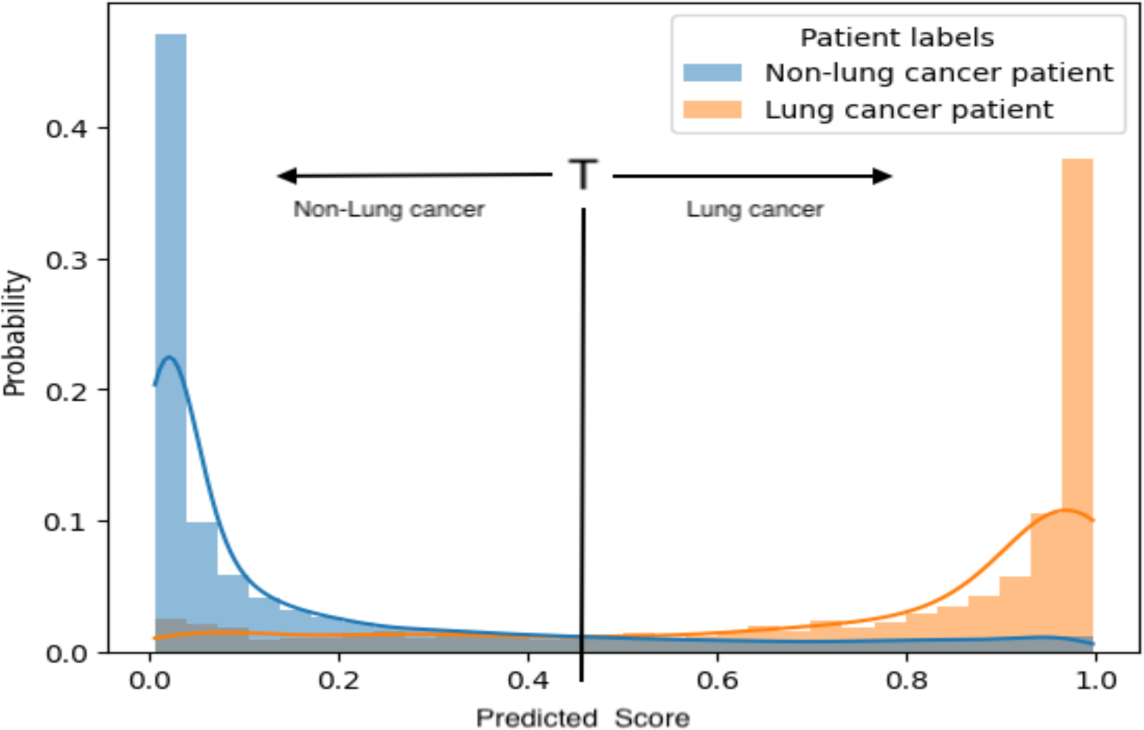
Prediction probability histogram of lung cancer prediction (normalised). T is the threshold between what is classified as Lung cancer and Non-Lung cancer

**Figure 5.**
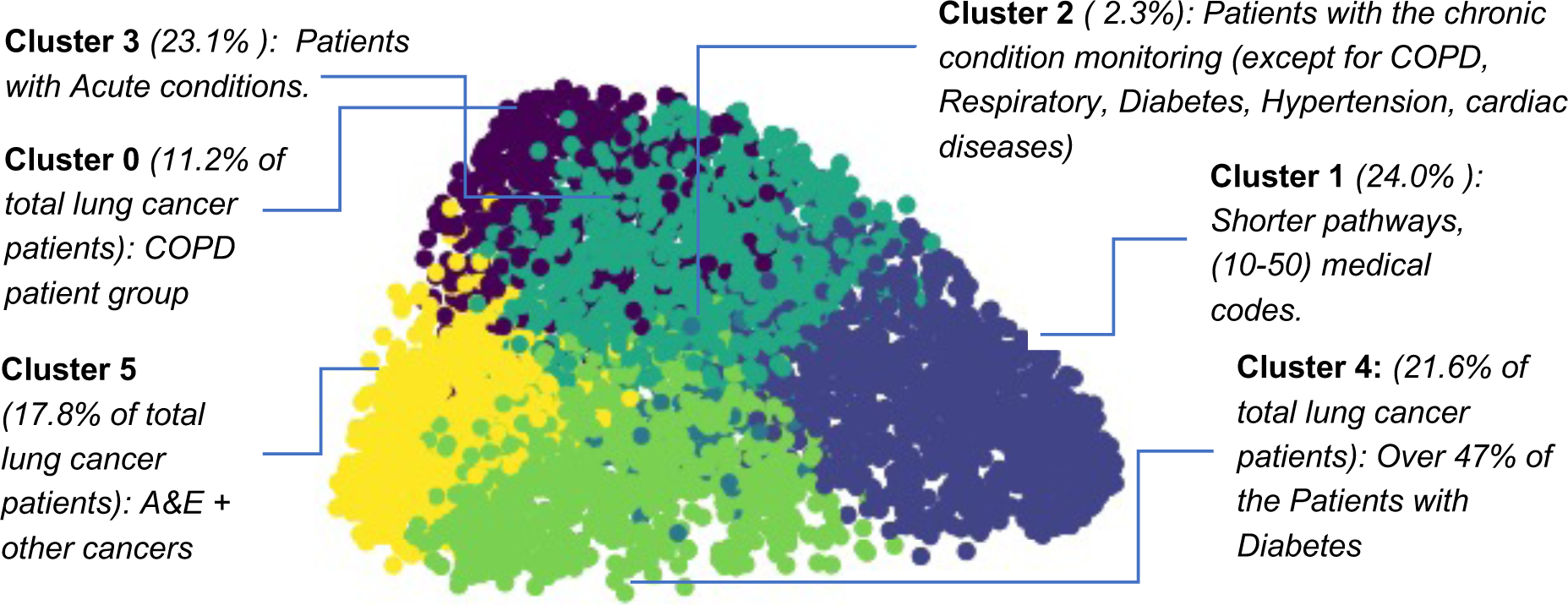
Six clusters obtained by clustering lung cancer patient pathways. A patient pathway representation is generated as a 768-dimensional vector by feeding the coded pathway to MedAlbert +LRC. To visualise the clustering of these pathways we use t-SNE algorithm for embedding high-dimensional data into a two-dimensional space.

**Table 3.** Predictive performance using three-year pathways at the chosen cut point. Models are trained on three-year pathways excluding the most recent one-month codes before diagnosis.

|  |  | **Precision**<br>(95% CI) | **Recall**<br>(95% CI) | **F1-score**<br>(95% CI) | **ROC-AUC**<br>(95% CI) |
| --- | --- | --- | --- | --- | --- |
| Logistic Regression Classifier (LR) | Not Lung Cancer | 99.8% (99.8–99.8) | 81.6% (81.6–81.7) | 89.8% (89.8–89.9) | 88.7% (88.4–88.9) |
|  | Lung Cancer | 3.1% (3.0–3.1) | 81.0% (80.4–81.6) | 5.9% (5.8–6.0) |  |
| MedAlbert + LRC | Not Lung Cancer | 99.9% (99.9–99.9) | 83.4% (83.3–83.5) | 90.9% (90.9–90.9) | 92.4% (92.1–92.7) |
|  | Lung Cancer | 3.6% (3.5–3.7) | 86.6% (85.3–87.8) | 6.8% (6.6–7.0) |  |

An additional factor is which cut point to take on the ROC curve to identify lung cancer patients. Table 4 shows the Sensitivity and PPV across the ROC. shows that the model separates lung cancer and not lung cancer populations well. In the UK, NICE accept a PPV of 3% as a threshold for fast-track investigation of suspected cancer. We therefore report the predictive performance of our selected model at the cut point of 0·4 in Table 4. The performance of the model by gender and ethnicity are reported in Table S11 and S12.

**Table 4.** Comparison of the prediction thresholds to identify patients with lung cancer diagnoses based on the validation cohort.

| Prediction Threshold | Number of patients with predicted lung cancer | Number of patients with correctly predicted lung cancer | Total number of Lung cancer patients in a validation dataset | Sensitivity | Positive predictive value |
| --- | --- | --- | --- | --- | --- |
| 0.4 | 6879 | 241 | 278 | 86.6% | 3.6% |
| 0.45 | 6145 | 234 | 278 | 84.0% | 3.8% |
| 0.5 | 5431 | 234 | 278 | 84.0% | 4.3% |
| 0.55 | 4799 | 221 | 278 | 79.4% | 4.6% |
| 0.6 | 4297 | 206 | 278 | 74.2% | 4.9% |
| 0.65 | 3758 | 210 | 278 | 75.7% | 5.6% |
| 0.7 | 3331 | 193 | 278 | 69.5% | 5.8% |

### Unsupervised learning of patient pathways reveals clinically relevant lung cancer patient groups

Clustering results are presented in **Error! Reference source not found.**. We chose the 6-cluster partition for a combination of robustness and separation of clinically relevant concepts. By computing the distribution of medical codes across lung cancer patients in each cluster (as shown in Figure S7 (left) – S12 (left) in Supplementary material), we can explore the patterns of the different clusters. In Cluster 0, Over 98% of patients are under COPD and codes related to chronic respiratory condition monitoring, whereas in Cluster 4 over 47% of patients have diabetes and 27% obesity. In Cluster 5 over 62% of the patients attended A&E and over 47% have another cancer while in Cluster 1, the three-year patient pathways contain relatively fewer (20–50) medical events/codes. The remaining two clusters have some overlaps with the above four. Cluster 2 only contains 2·4% of the cohort and all of them are under chronic condition monitoring (except for COPD, Respiratory, Diabetes, Hypertension, cardiac diseases) while most patients in Cluster 3 present acute conditions.

## Discussion

Although population-based lung cancer risk models have been built using machine learning techniques such as random forests and vector boost, these are static models that do not account for temporal relationships between data elements in EHR data. In addition, population models of risk and predictive models for symptomatic patients are different and serve different clinical purposes. We focus on the latter here. Our model for lung cancer early detection, based on MedAlbert plus a LRC achieved an AUROC of 0·924(0·921, 0·927) with a Sensitivity of 86·6%, Specificity 83·4%, PPV 3·6%, and NPV 99·9% based on the three year’s data prior to diagnosis less the one immediate month before. The current specific clinical model, QCancer Lung, has a PPV of 1.34% at its maximum sensitivity of 77.3%. ^8^ Capturing the subtle differences in presentations between cancer and non-cancer pathways to diagnosis enables much more accurate models. As far as we are aware this is the first time ALBERT has been used to analyse EHR data coded in a rich terminology such as Read or SNOMED-CT. A recent publication has used NLP to extract clinical concepts from unstructured text EHR data in the US, but the final analysis was based on a multivariable regression model. ^24^ In that study finger clubbing, cough, haemoptysis, wheeze, weight loss, back pain, bone pain, shortness of breath, and fatigue were significant predictors of lung cancer but the model was not validated to provide an estimate of performance. Using MedAlbert for structured data enables the subtle differences in presentations between cancer and non-cancer pathways to diagnosis to be captured. Using cluster analysis this can be interpreted in terms of what clinical concepts the model is fixing its ‘attention’ to, indeed, on account of fine tuning with a LRC explainability at individual patient level is also possible if the attention scores are extracted. Symptoms picked out in the clusters include breathlessness, chest pain, haemoptysis, cough, and ‘general symptoms’ (including in our analysis weakness and malaise). In addition, a number of ENT, lower GI, and musculoskeletal symptoms appear in the attention scores. This may be as they are either associated but not causal, or temporally related in the patient’s care pathway. The cluster analysis shows other known risk factors; COPD and other respiratory conditions, age, gender, and smoking history. Diabetes is known to be associated with a number of cancers, particularly liver and pancreas but hasn’t been associated with lung cancer previously. ^25,26^ Obesity on the other hand has previously been found to be associated with a lower risk of lung cancer, but the diabetes cluster shows obesity as a risk factor with the highest incident rate. It may be the strong association with diabetes that is bringing it into this cluster. ^27^ Alcohol has been linked with head and neck and upper GI cancers in particular but a recent study also suggests that lung cancer risk may also be increased, possibly by genetic differences in acetaldehyde production.^28^ The association of other cancers with lung cancer are shown in cluster 5, specifically oral cancer, colorectal cancer, breast cancer, uterine cancer, cervical cancer, renal cancer, ovary cancer, prostate cancer and gastro-oesophageal cancer (Figure S13).

A ‘Pathway to Diagnosis’ for a patient, as defined in our study, contains the most possible elaboration of the coded medical records of each patient. It consists of as many types of medical codes as possible (such as symptoms, diagnoses, medications, procedures, sites of encounter, and medical tests) appearing during three years so that it possesses a wealth of information of disease progression from the perspective of patients and clinical investigation process from the perspective of clinicians and stands in contrast to most other statistical methods where data has to be aggregated, reducing its dimensionality to enable analysis. We view the pathways as a medical language for describing patients’ health details and medical experience where the vocabulary is all the unique medical codes that make up each pathway and the grammar is how the codes relate to each other in the context of each pathway. We propose a novel model, MedAlbert, based on the state-of-the-art NLP techniques for learning deep pathway representations from large amounts of EHRs that capture rich medical code relationships and dependencies. This allows us to discover lung cancer progression patterns and clinical investigation patterns, as well as the associations of patient pathways with the underlying health status of patients and the corresponding diagnoses.

Removing the one month of data before diagnosis may reduce the potential for ‘red flag’ symptoms to be picked up the model. In future, prospective data capture from primary care should enable ‘index consultations’ to be identified prior to cancer diagnosis excluding the impact of post-referral events. This better construction of the source records,^29^ along with direct linkage of secondary care data would enable distinguishing bias from signal in the immediate pre-diagnosis period. We took a conservative approach in excluding the one month prior to diagnosis in this study which may have underestimated the accuracy of the model. The observed lack of symptoms in the clusters and attention factors most likely reflect a lack of coding of symptoms and signs in UK primary care EHR data and additional means of coding these, or use of NLP to extract clinical concepts from text should be employed in future work.

Although MedAlbert-based models of prediction using coded clinical pathways appear to have good validity and appear to be based around concepts with support in the medical literature, their use in clinical practice is constrained at present by several factors. Firstly, the model needs to be validated in an external clinical dataset rather than a 30% partition of the starting data. Secondly, the implementation of a model containing potentially most of the data points in a three-year patient history might be difficult to achieve technically in real time in consultations unless EHR data is pre-processed. Methods for improving the performance of HL7 FHIR Application Programming Interfaces for extracting large medical records quickly or enabling selective export as well as exploring local-ICT constraints on processing are required. However, the future of diagnosis in primary care will lie in the operation of AI supported clinical diagnosis and the technology and EHR systems will have to adapt to support that.^30^ The approach taken can be extended to predict other cancers and other diagnoses to provide a generic diagnostic support for primary care, however, methods for combining the non-independent risks of the potential diagnoses will have to be applied.

MedAlbert is not a Large Language Model (LLM) in that it is many fold smaller than the models that have become commercially available since early 2022, proving to be a potentially powerful tool in NLP in particular. Further work in the area should explore to what extent a pre-trained LLM is a more powerful tool for the approach adopted here compared with a pretrained ALBERT model, or simply an additional overhead. The area of explainability and computational representation of explainability is a key area of research in diagnostic AI. Our approach offers individual patient level explainability, in addition to interpretability at population level that a general LLM does not. Trust and uptake of AI models in clinical settings are heavily influenced by explainability and much further work on learning and modelling predictions of the MedAlbert model to provide patient-level constructs such as knowledge graphs to drive explanations in the EHR is needed. In addition, LLMs are not currently able to be authorised as medical devices and their use is restricted to areas that can be claimed as not ‘directly’ influencing clinical care. Prediction of possible clinical diagnoses during a primary care encounter is without doubt a medical device, and our approach illustrates how the analytical power of transformers can be leveraged without running into the legal, ethical and regulatory issues posed by LLM such as ChatGPT.

## Ethics

Ethical approval was from London Bromley Research Ethics Committee ID: 252487 REC Reference: 18/LO/2240. Data Access was approved by the WSIC Data Access Committee. All data used in this paper were fully anonymized before analysis.

## Contributions of authors

LW conducted data analysis and model building, developing, and validation, and wrote first draft of the manuscript. YY jointly designed and developed the models with Lan. MB contributed to the design of the study and supervised data analysis methods and model development. RP developed pathway clustering and visualisation. BG contributed to the data access and interpretation, analysis and interpretation of the results. BD obtained funding and contributed to the conceptualisation and design of the study. BD also conducted code dimension reduction, clinical interpretation, iteration of design and drafting of the manuscript. EK contributed to the conceptualisation and design of the study and helped with data access, clinical interpretation, and iteration of design. BD and ER are Joint senior authors. All authors contributed to drafting of the manuscript.

## Data Availability

Due to data governance limitations, the deidentified patient data used to develop and validate the models cannot be shared.

## Acknowledgements

We thank Eamon O’Doherty (Northwest London Clinical Commissioning Group, London, UK) for writing SQL scripts for deriving lung cancer patient pathways to diagnosis and for correcting the data we need to use in our study as well as data access and analysis guidance.

## Data sharing

Due to data governance limitations, the deidentified patient data used to develop and validate the models cannot be shared. The model code will be available on GitHub at https://github.com/Lung_cancer_prediction/MedAlbert.

