## Supplementary material for "Transformer-based deep learning model for the diagnosis of suspected lung cancer in primary care based on electronic health record data"

### Methods

#### Defining patient level pathways to diagnosis

Lung cancer patients were identified from the data using codes in Table S*1*.

| | |
|---|---|
| B22z.; Byu20 | Malignant neoplasm of bronchus or lung NOS |
| B22.. | Malignant neoplasm of trachea, bronchus and lung |
| B2211 | Malignant neoplasm of hilus of lung |
| B222. | Malignant neoplasm of upper lobe, bronchus or lung |
| B2221 | Malignant neoplasm of upper lobe of lung |
| B222z | Malignant neoplasm of upper lobe, bronchus or lung |
| B224. | Malignant neoplasm of lower lobe, bronchus or lung |
| B2241 | Malignant neoplasm of lower lobe of lung |
| B224z | Malignant neoplasm of lower lobe, bronchus, or lung |
| B223. | Malignant neoplasm of middle lobe, bronchus or lung |
| B2231 | Malignant neoplasm of middle lobe of lung |
| B223z | Malignant neoplasm of middle lobe, bronchus or lung |
| B225. | Malignant neoplasm of overlapping lesion of bronchus & lung |
| B22y. | Malignant neoplasm of other sites of bronchus or lung |
| B570. | Secondary malignant neoplasm of lung |

Table S1 Read Codes CT v2 for Lung Cancer

#### Curating and grouping medical codes

Each medical code appearing in a patient's clinical records is a feature of the pathway to diagnosis. Whether we use the exact medical codes or group the codes, and the granularity of code groups determines the resolution of each pathway. The use of various levels of granularity of medical groups allows us to preserve coarse-grained or fine-grained details in each pathway for further deep representation learning according to our analytical requirement.

High input resolution (i.e. using the exact 31,312 collected original medical codes) can largely improve the accuracy of a model during the training process. However, high resolution brings more parameters to the model and hence the complexity of the model rises. Without selecting and grouping, the model must learn from too many features. In particular, the varied presence of many minor features and noisy information (e.g. administration info) in the training dataset can reduce model prediction. As more and more parameters are added to a model and the complexity of the model exceeds a certain point, we risk over-fitting our model and the model may lack generalizability.

On the contrary, low input resolution by selecting and grouping medical codes with prior knowledge from medical experts reduces the parameters and hence the complexity of the model. The variance falls when applying the model to real-world data. However, prior knowledge related to cancer is limited and may cause the model to deviate from the accurate representation of real-world data. Relying too much on prior knowledge, for example likely symptom codes or known risk factors, also limits the ability of the model to discover unknown patterns.

Therefore, we need to find a balance in our representation model. It is the level of complexity at which the sacrifice of accuracy is equivalent to the reduction in variance. It is necessary to have input from medical experts, based on which we will select, and group medical codes and pre-train a pathway representation model using grouped and original medical codes as input respectively. However, care must be taken not to introduce bias, so the strategy of applying the same selection rules with respect to granularity to all code groups was adopted. Based on the results by applying the learned representations (features) on some clustering and prediction tasks, we can discover how relevant the medical codes (groups) are to lung cancer diagnosis and optimise the model with the appropriate number of input medical codes (groups).

Read codes were used in WSIC at the time of our study and they have the advantage of being organised hierarchically, that is moving from more general terms (e.g. 'H: Respiratory system diseases') to more specific terms (e.g. 'H06z0:Chest infection') as you move down the hierarchy, which makes it easier to group medical codes and define the medical code groups at different levels of granularity. For example, the first character corresponds to the first level group. If we need a bit finer group, we can define a second level code group by aggregating the codes beginning with some specified two characters. This process can be done with SNOMED codes but only within individual SNOMED hierarchies and with the support of a terminology service to map the terms.

We curated and grouped the Read codes based on both prior knowledge from medical experts and the usage frequency of each medical code (group) among patients.

We first sorted the collected Read codes by the order from '0' to '9' and then 'A' to 'Z'. We then select and group codes following the read hierarchy, from more general terms to more specific terms.

Criteria of selecting and grouping medical codes

1. Based on the advice from the medical expert, we select the Read codes regarding disease progression such as the symptoms and signs, diagnoses, lifestyle and medical history, as well as the codes revealing disease investigation processes such as the diagnostic tests, procedures, medications and sites of encounter. We excluded the Read codes used for recording administrative procedures. For example, the read codes beginning with '91', '92', '93' about patient registration and records and some codes about 'letter send' and 'email send'. The read codes beginning with '41' related to 'Laboratory procedures - general', provide administrative information as the codes from '42'…' 'o '4Q'..' have provided sufficient information for recording specific Laboratory tests. We encoded the fact that a test had been carried out, not its result.
2. Analysis of usage frequency of each medical code (group) among patients. As shown in Table S*2*, only a small portion (0·02%) of Read codes are presented in the records of the majority (80% and over) of patients. Only 9·62% of the total collected codes are used by over 1% of the patients while 65·31% of the codes are used by fewer than 0·1% of the patients. The expected (mean) usage frequency of the read codes is 0·87% of the total number of the patients. Therefore, we set a threshold at 1%. If a medical code (group) appears in the records of more than 1% of the total lung cancer patients or non-lung cancer patients, we will consider picking these codes out and aggregating them into a separate group. In this way, the resulting total number of code groups will not exceed 2000.

| Proportion of read codes | 0·02% | 0·2% | 0·3% | 0·48% | 1·26% | 9·62% | 84·83% | 65·31% |
|---|---|---|---|---|---|---|---|---|
| **Proportion of lung cancer patients** | 80% and over | 70% and over | 50% and over | 30% and over | 10% and over | 1% and over | 0·5% and under | 0·1% and under |

Table S2: Usage frequency of lung cancer related read codes

3. Combining read code groups. We combined the read code groups beginning with different characters based on our analytical requirements. For example, as read codes beginning with '196..'and 'R09' are all about "Abdominal pain", the two read code groups can be combined.

### Machine learning approach

#### Logistic Regression Model (LR) comparator

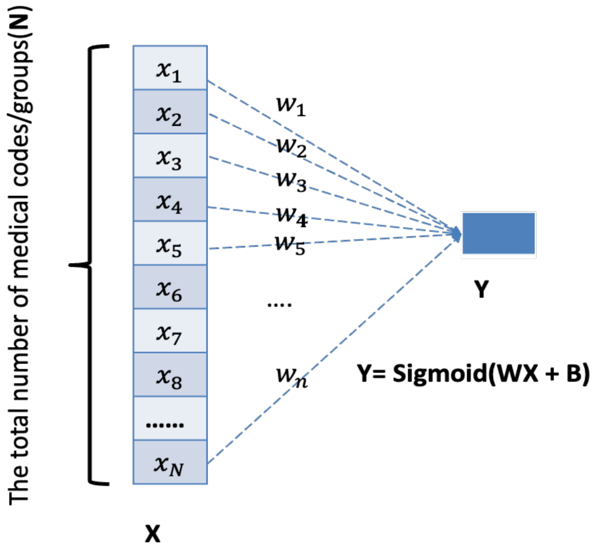


Figure S1 The architecture of the Logistic Regression Classifier for lung cancer prediction. The input for a given patient is a binary vector indicating the presence of a medical code across the entire patient pathway. This model inherently ignores the sequence of medical codes across a patient's pathway.

To create a single Logistic Regression (LR) classifier for lung cancer prediction as a comparator model, we represented the pathway of each patient as a vector of size $N$ and used it as the input of the model. $N$ is the total number of medical codes/groups ($N = 450$ in our study) we used to derive patient pathways excluding the lung cancer diagnostic codes. Each entry of the vector corresponds to a medical code/group. $x_i = 1$, if the medical code/group $i$ is present in the pathway, otherwise, $x_i = 0$. $w_i$ $(i = 1,2,\ldots,N)$ is the contribution/weight of each medical code/group to lung cancer prediction.

### Results

In the period from January 1981 to December 2020 there were in total 3,303,992 patients in WSIC, where 11,847 were diagnosed with lung cancer. The number of lung cancer diagnoses per month between January 1981 and December 2020 is shown in Figure S*2*. Figure S*3* shows how we construct training and validation datasets.

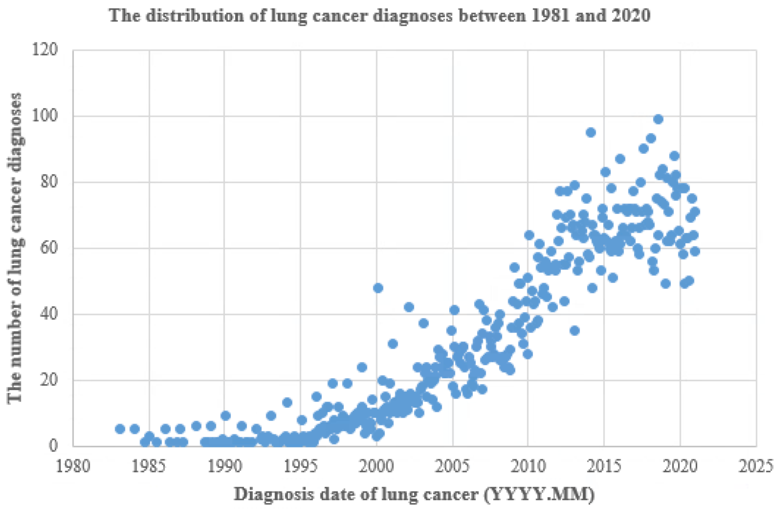


Figure S2 The number of patients diagnosed with lung cancer per month between Jan 1981 and Dec 2020.

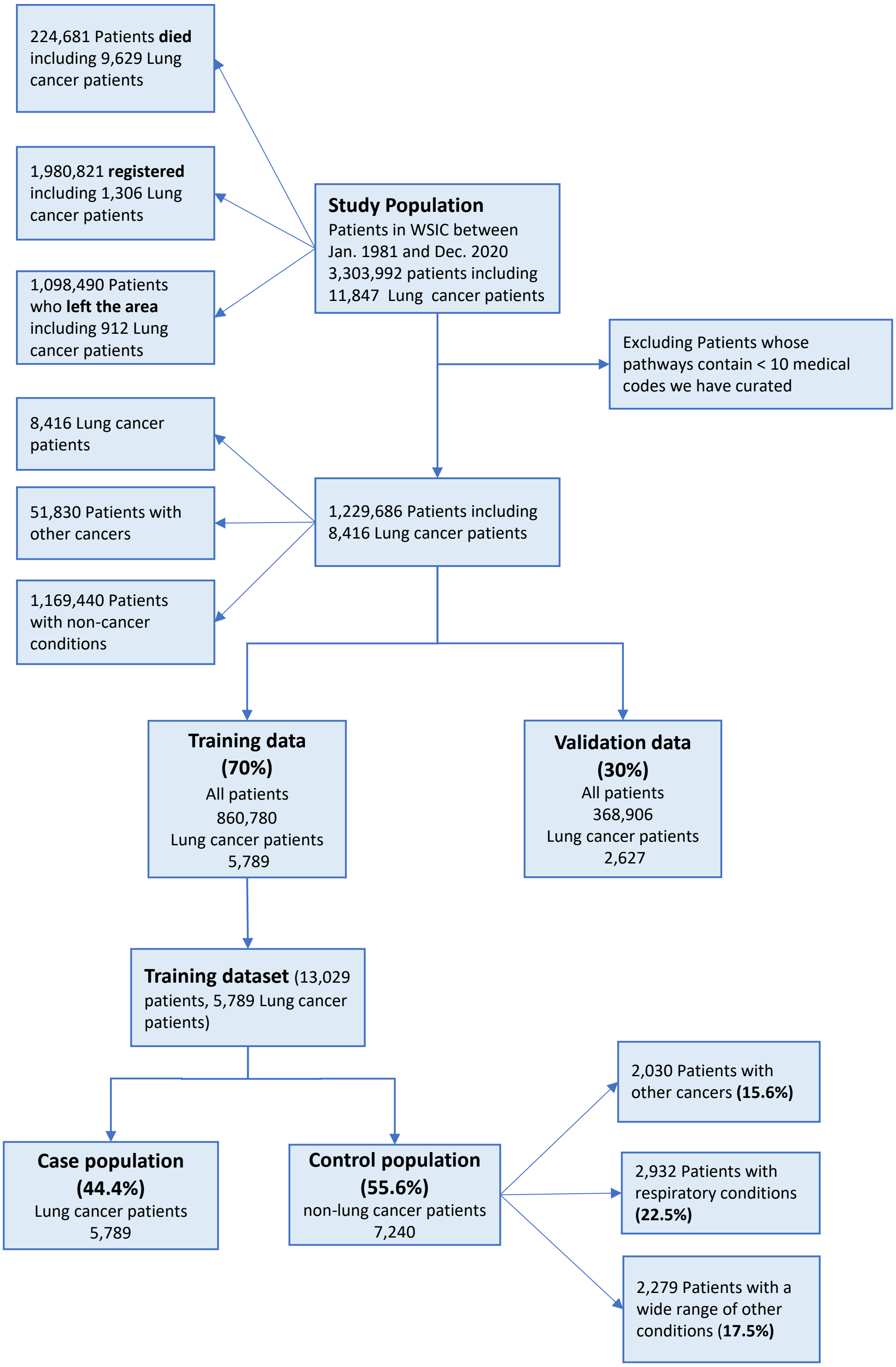


Figure S3 Constructing training and validation datasets.

**Sensitivity analysis of one- and two-year pathways prior to diagnosis**

For early diagnosis of one and two years, MedAlbert+LRC still outperforms the single LR classifier, obtaining 1%–9% improvements in Precision, Recall, F1 score and ROC-AUC ( Table S*3* – Table S*10*) . The very low precision is because the ratio of non-lung cancer patients and lung cancer patients in the validation dataset is 50: 1 (being the incidence of Lung Cancer in NW London), which causes much higher False Positive compared with True Positive. The results are reported with 95% confidence interval (CI).

| | | **Precision** | **Recall** | **F1-score** | **ROC-AUC** |
|---|---|---|---|---|---|
| Logistic Regression Classifier (LR) | Not Lung Cancer | 99·8% (99·8, 99·8) | 90·8% (90·8, 90·9) | 95·1% (95·1, 95·1) | 89·6% (89·3, 89·9) |
| | Lung Cancer | 5·1% (5·0, 5·2) | 69·4% (68·4, 70·5) | 9·5% (9·2, 9·7) | |
| MedAlbert + LRC | Not Lung Cancer | 99·9% (99·9, 99·9) | 93·6% (93·5, 93·8) | 96·7% (96·5, 96·8) | 96·8% (96·6, 97·0) |
| | Lung Cancer | 9·0% (8·8, 9·1) | 88·1% (87·5, 88·7) | 16·2% (16·0, 16·5) | |

Table S3: Predictive performance on the three-year pathways. (Models are trained on full three-year pathways)

| | | **Precision** | **Recall** | **F1-score** | **ROC-AUC** |
|---|---|---|---|---|---|
| Logistic Regression Classifier (LR) | Not Lung Cancer | 99·8% (99·7, 99·8) | 82·2% (82·1, 82·3) | 90·1% (90·1, 90·2) | 86·2% (85·8, 86·5) |
| | Lung Cancer | 2·8% (2·8, 2.9) | 73·9% (73·0, 74·8) | 5·4% (5·3, 5·6) | |
| MedAlbert + LRC | Not Lung Cancer | 99·9% (99·8, 99·9) | 86·4% (86·3, 86·5) | 92·6% (92·6, 92·6) | 89·9% (88·8, 91·0) |
| | Lung Cancer | 2·9% (2·8, 3·0) | 75·9% (72·6, 79·2) | 5·6% (5·4, 5·8) | |

Table S4: Predictive performance on the three-year pathways· (Models are trained on three-year pathways excluding the most recent three-month read codes before diagnosis)

| | | Precision | Recall | F1-score | ROC-AUC |
|---|---|---|---|---|---|
| Logistic Regression Classifier (LR) | Not Lung Cancer | 99·6% (99·6, 99·6) | 89·7% (89·7, 89·8)) | 94·4% (94·4, 94·4) | 80·5% (80·1, 80·9) |
| | Lung Cancer | 3·2% (3·1, 3·3) | 47·4% (46·4, 48·3) | 6·0% (5·8, 6·1) | |
| MedAlbert + LRC | Not Lung Cancer | 99·8% (99·7, 99·8) | 57·6% (57·4, 57·7) | 73·0% (72·9, 73·1)) | 72·3% (70·8, 73·8) |
| | Lung Cancer | 0·9% (0·9, 1·0) | 75·5% (72·8, 78·2) | 1·9% (1·8, 1·9) | |

Table S5: Predictive performance on the two-year pathways occurring one year earlier than diagnosis· (Models are trained on full three-year pathways)

| | | Precision | Recall | F1-score | ROC-AUC |
|---|---|---|---|---|---|
| Logistic Regression Classifier (LR) | Not Lung Cancer | 99·6% (99·6, 99·6) | 88·1% (88·0, 88·1) | 93·3 % (93·3, 93·3) | 83·0% (82·7, 83·3) |
| | Lung Cancer | 2·8% (2·7, 2·9) | 56·3% (55·5, 57·2) | 6·2% (6·0, 6·3) | |
| MedAlbert + LRC | Not Lung Cancer | 99·7% (99·7, 99·8% | 89·5% (89·4, 89·6) | 94·4% (94·3, 94·4) | 86·3% (85·3, 87·2) |
| | Lung Cancer | 2·9% (2·7, 3·0) | 57·1% (53·6, 60·7) | 5·4% (5·1, 5·8) | |

Table S6: Predictive performance on the two-year pathways occurring one year earlier than diagnosis. (Models are trained on three-year pathways excluding the most recent one-month codes before diagnosis)

| | | Precision | Recall | F1-score | ROC-AUC |
|---|---|---|---|---|---|
| Logistic Regression Classifier (LR) | Not Lung Cancer | 99·7% (99·7, 99·7) | 79·9% (79·8,79·9) | 88·6% (88·6, 88·7) | 82·5% (82·1, 82·8) |
| | Lung Cancer | 2·3% (2·3, 2·4) | 68·8% (67·7, 69·4) | 4·9% (4·8, 5·0) | |
| MedAlbert + LRC | Not Lung Cancer | 99·8% (99·8, 99·8) | 84·7% (84·6, 84·8) | 91·6% (91·6, 91·7) | 86·6% (85·2, 88·0) |
| | Lung Cancer | 2·4% (2·3, 2·5) | 69·8% (66·3, 73·4) | 4·6% (4·4, 4·9) | |

Table S7: Predictive performance on the two-year pathways occurring one year earlier than diagnosis. (Models are trained on three-year pathways excluding the most recent three-month read codes before diagnosis)

| | | Precision | Recall | F1-score | ROC-AUC |
|---|---|---|---|---|---|
| Logistic Regression Classifier (LR) | Not Lung Cancer | 99·4% (99·4, 99·5) | 88·2% (88·2, 88·2) | 93·5% (93·5, 93·5) | 76·6% (76·4, 76·9) |
| | Lung Cancer | 3% (3, 3·1) | 42·5 %(41·9, 43·1) | 5·6% (5·5, 5·7) | |
| MedAlbert + LRC | Not Lung Cancer | 99·8% (99·7, 99·8) | 50·6% (50·4, 50·7) | 67·1% (67·0, 67·3) | 68·8% (67·5, 70·1) |
| | Lung Cancer | 0·8% (0·8, 0·9) | 77·5% (75·3, 79·8) | 1·7% (1·6, 1·7) | |

Table S8: Predictive performance on the one-year pathways occurring two years earlier than diagnosis. (Models are trained on full three-year pathways)

| | | Precision | Recall | F1-score | ROC-AUC |
|---|---|---|---|---|---|
| Logistic Regression Classifier (LR) | Not Lung Cancer | 99·5% (99·5, 99·5) | 86·5% (86·4, 86·5) | 92·5% (92·5, 92·5) | 80·3% (80·1, 80·5) |
| | Lung Cancer | 3·1% (3·2, 3·3) | 47·4% (46·8, 48·4) | 5·8% (5·8, 5·9) | |
| MedAlbert + LRC | Not Lung Cancer | 99·8% (99·7, 99·8) | 92·5% (92·4, 92·6) | 96·0% (95·9, 96·0) | 83·3% (81·8, 84·7) |
| | Lung Cancer | 3·2% (3·1, 3·3) | 52·4% (49·3, 55·4) | 6·0% (5·8, 6·2) | |

Table S9: Predictive performance on the one-year pathways occurring two years earlier than diagnosis. (Models are trained on three-year pathways excluding the most recent one-month read codes before diagnosis)

| | | Precision | Recall | F1-score | ROC-AUC |
|---|---|---|---|---|---|
| Logistic Regression Classifier (LR) | Not Lung Cancer | 99·6% (99·6, 99·7) | 76·9% (76·9, 76·7) | 86·8% (86·7, 86·9) | 79.3% (78·9, 80·1) |
| | Lung Cancer | 2·7% (2·6, 2·9) | 67·0% (65·5, 70·7) | 5·1% (4·9, 5·3) | |
| MedAlbert + LRC | Not Lung Cancer | 99·7% (99·7, 99·8) | 88·1% (88·0, 88·2) | 93·6% (93·5, 93·7) | 83·5% (82·2, 85·4) |
| | Lung Cancer | 2·4% (2·3, 2·5) | 55·4% (52·4, 58·5) | 4·7% (4·5, 4.9) | |

Table S10: Predictive performance on the one-year pathways occurring two years earlier than diagnosis. (Models are trained on three-year pathways excluding the most recent three-month read codes before diagnosis)

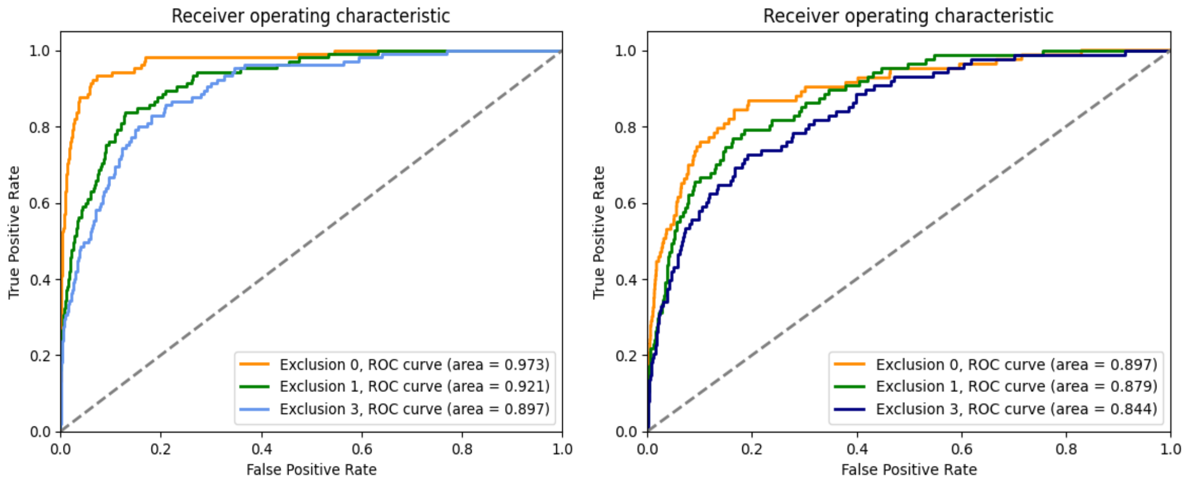


Figure S4: ROC curve of MedAlbert+ LRC (left) and a single LR classifier (right) applied on three-year patient pathways before diagnosis

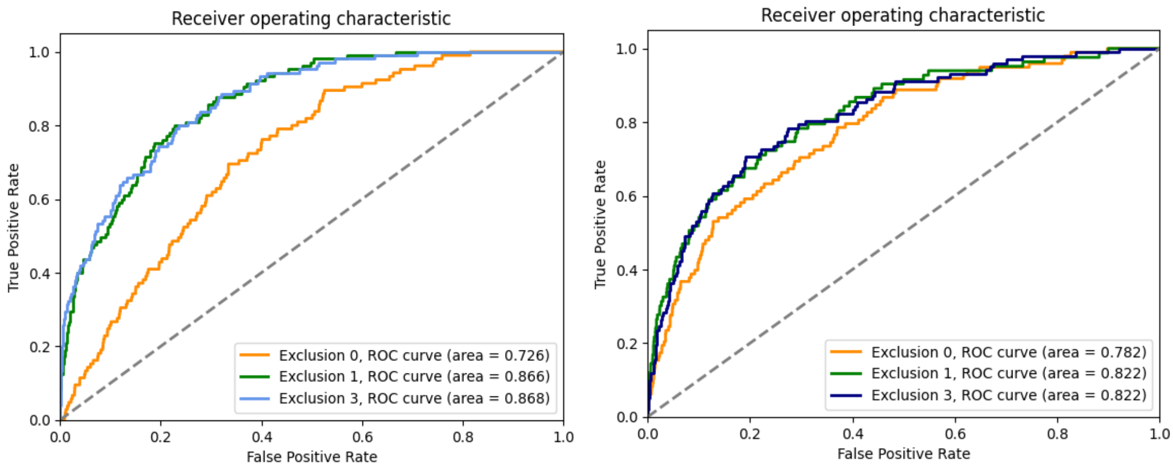


Figure S5: ROC curve of MedAlbert+ LRC (left) and a single LR classifier (right) on the two-year pathways occurring one year earlier than diagnosis.

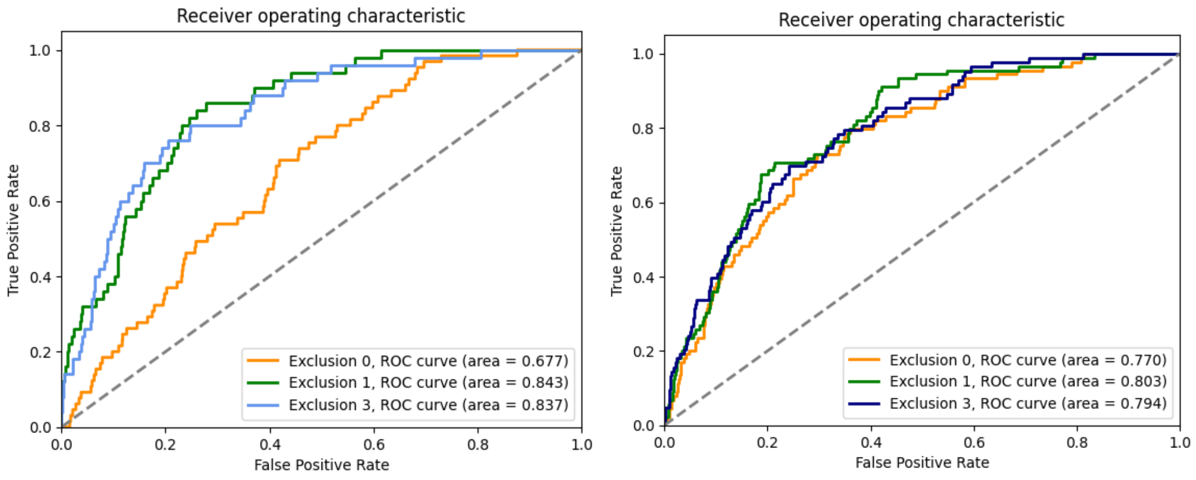


Figure S6: ROC curve of MedAlbert+ LRC (left) and a single LR classifier (right) on the one-year pathways occurring two years earlier than diagnosis.

| Gender | | Precision | Recall | F1-score | ROC-AUC |
|---|---|---|---|---|---|
| Female | Not Lung Cancer | 99·9% (99·9, 99·9) | 88·4% (88·3, 88·5) | 93·8% (93·7, 93·8) | 93·6% (93·2, 94·1) |
| | Lung Cancer | 3·7% (3·5, 3·9) | 83·1% (80·9, 85·4) | 7·1% (6·7, 7·5) | |
| Male | Not Lung Cancer | 99·9% (99·8, 99·9) | 75·7% (75·6, 75·8) | 86·1% (86·0, 86·2) | 91·2% (91·0, 91·4) |
| | Lung Cancer | 1·9% (1·8, 2·1) | 89·2% (87·0, 91·5) | 3·8% (3·6, 3·9) | |

Table S11 Predictive performance of the MedAlbert + LRC by gender. (Models are trained on three-year pathways excluding the most recent one-month read codes before diagnosis)

| Ethnicity | AUROC | Ethnicity | AUROC | Ethnicity | AUROC |
|---|---|---|---|---|---|
| British | 90·9% (88·9, 92·0) | Indian | 88·1% (87·1, 90·2) | African | 93·7% (93·5, 94·1) |
| Irish | 91·2% (89·7, 92·6) | Pakistani | 89·8% (89·1, 90·9) | Caribbean | 90·6% (89·6, 91·7) |
| Other White | 93·4% (92·6, 94·2) | Chinese | 86·6% (86·1, 87·3) | Other Asian | 91·0% (90·0, 92·1) |
| Other | 92·9% (91·7, 94·0) | Bangladeshi | 94·7% (94·2, 95·3) | | |

Table S12: AUROC of the MedAlbert + LRC by ethnicity (Models are trained on three-year pathways excluding the most recent one-month read codes before diagnosis)

**Unsupervised learning of patient pathways reveals clinically relevant lung patient groups**

We present the distribution of medical codes across patient pathways in each cluster (Figure S7 (left) – Figure S12 (left)) and the distribution of medical codes which the predictive model mostly attends to across patient pathways in each cluster (Figure S7 (right) – Figure S12 (right))

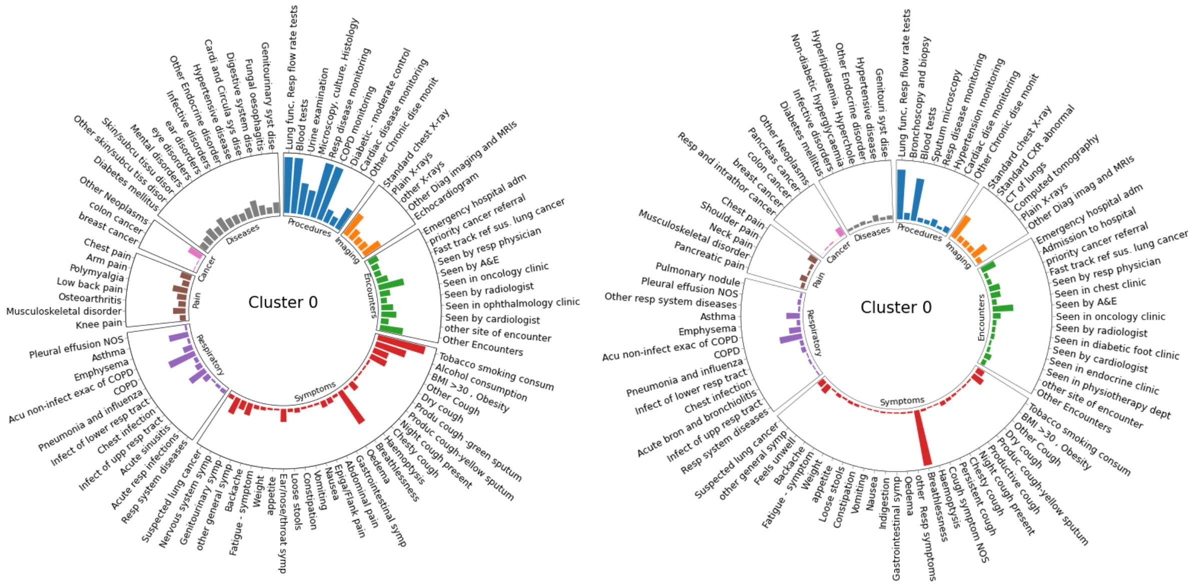


Figure S7 Cluster 0 (11·2% of total lung cancer patients): over 98% of Lung cancer patients with COPD and chronic respiratory conditions.

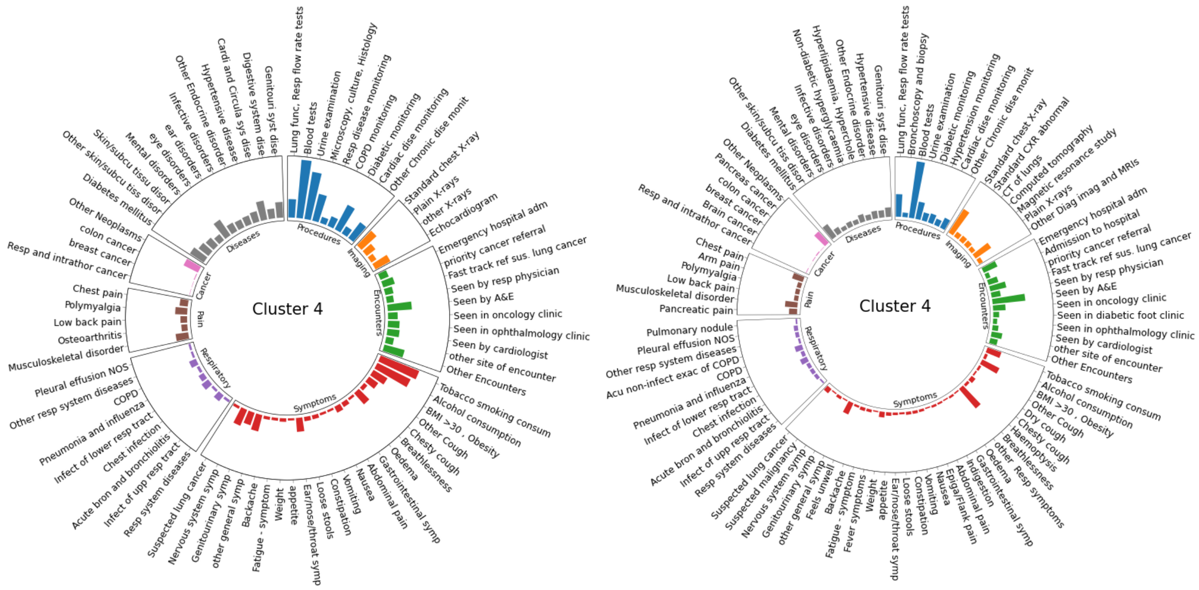


Figure S8 Cluster 4 (21·6% of total lung cancer patients): Over 47% of lung cancer patients have diabetes, over 27% have Obesity

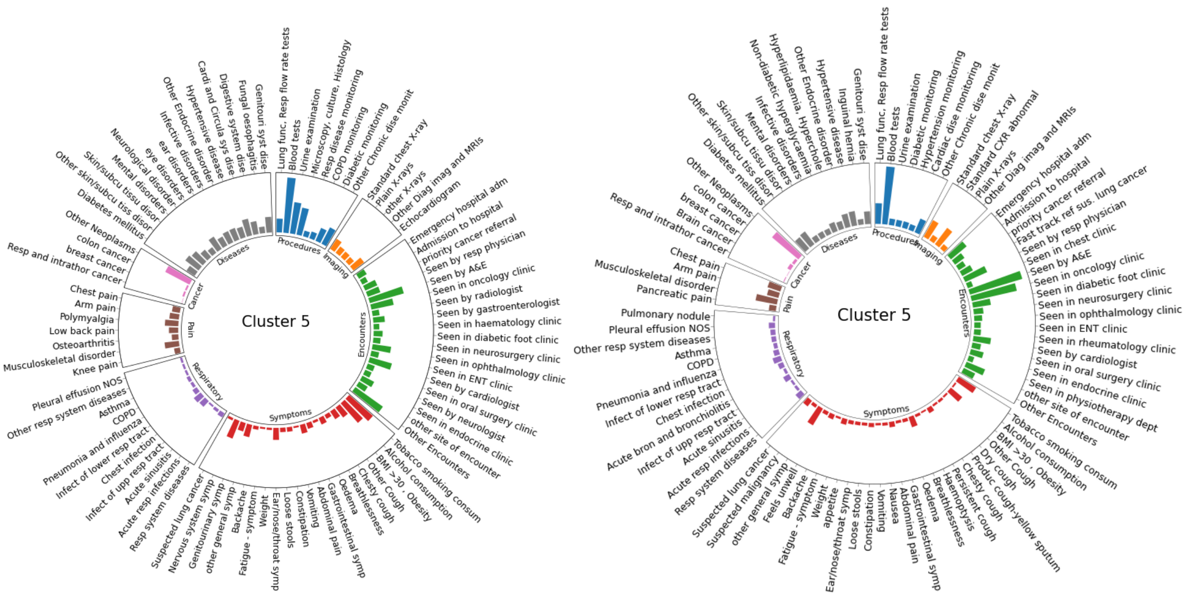


Figure S9 Cluster 5 (17·8% of total lung cancer patients): Over 62% of lung cancer patients attended A&E, over 47% have another cancer

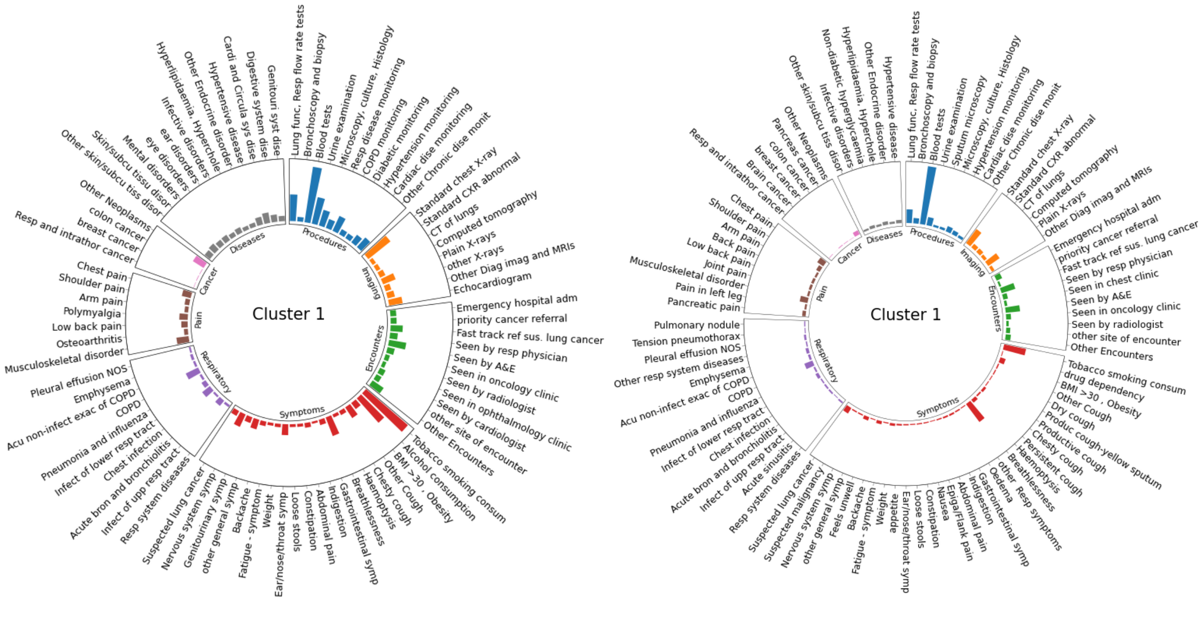


Figure S10: Cluster 1 (24·0% of total lung cancer patients) : the three-year pathways contain much fewer (10 – 50) medical codes than other clusters.

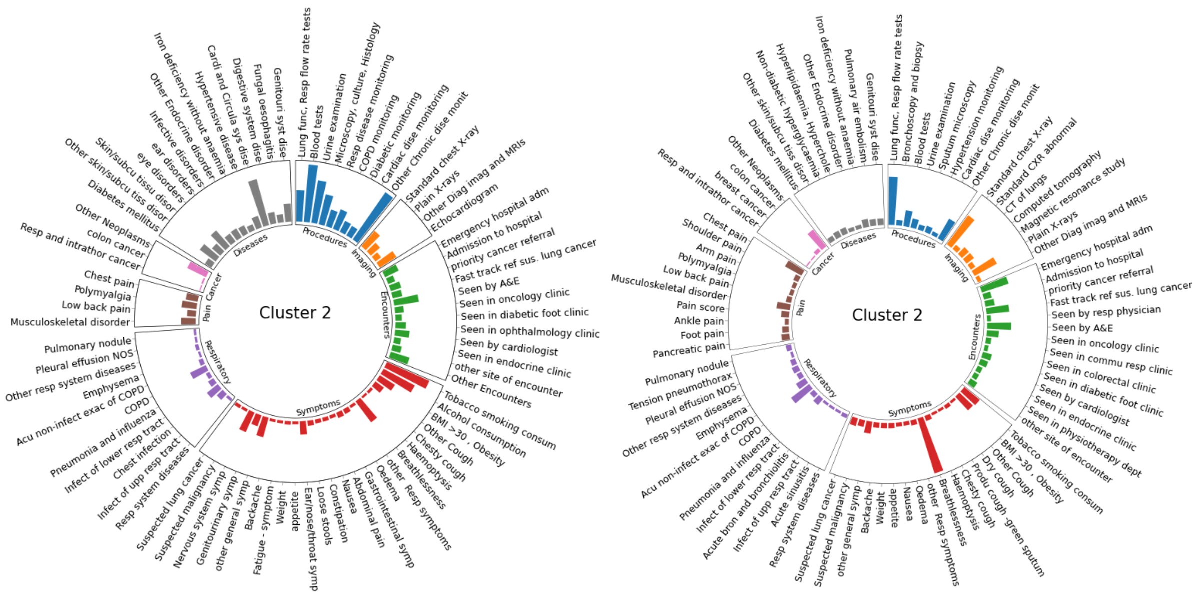


Figure S11: Cluster 2 (2·3% of total lung cancer patients): 100% of this cluster's patients are under the chronic condition monitoring (except for COPD, Respiratory, Diabetes, Hypertension, cardiac diseases)

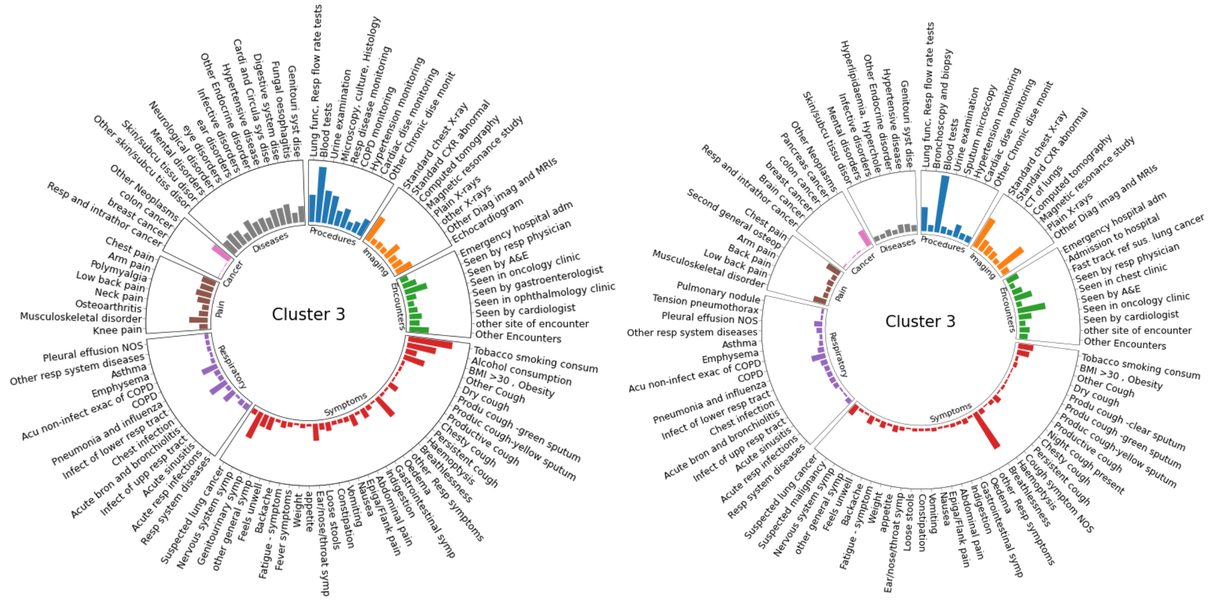


Figure S12: Cluster3 (23·1% of total lung cancer patients): Most patients in this cluster are with acute conditions·

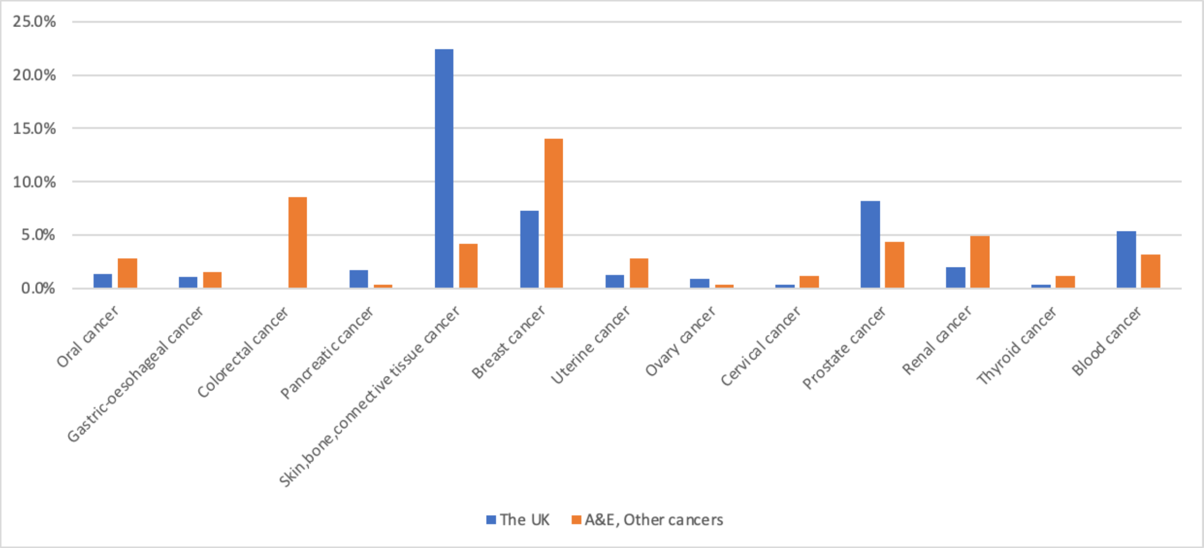


Figure S13: In Cluster 5: A&E + other cancers, the distribution of lung cancer patients who were also diagnosed with other cancers. The cancer incidences are compared with that in the UK

As shown in Figure S7 - Figure S*13*, in the cluster of A&E+other cancers (17·7% of total lung cancer patients), lung cancer patients are featured by A&E attendances(62·8%), oncology clinic encounters (40·2%) and the diagnosis of other cancers (47·9%) before lung cancer diagnosis· We list the cancers they suffered from in Figure S*13*. We can see the lung cancer patients in this cluster had much higher cancer incidences of Oral (2·8%), Gastric-oesophageal (1·5%), Colorectal (8·5%), Breast (14%), Uterine (2·8%), Renal (4·9%), Cervical (1·1%), Thyroid (1·1%) than those in the population of the UK. We would like to discuss the association of lung cancer with the other cancers, Musculoskeletal disorder, disorder of spine, Back disorders (N…), Cardiac and circulatory system diseases, skin and subcutaneous tissue disorders (M…), eye conditions and Diabetes as well as encounters, such as A&E, lung function test, plain X-ray, CXR, seen by cardiologist.

In the Diabetes cluster (21·6% of total lung cancer patients), except for Diabetes and Obesity, around 30% of the lung cancer patients also suffered from Cardiac and circulatory system diseases, eye conditions, other cancers and Genito-urinary system diseases. This cluster contains the highest proportion of drinkers (63·9%). Our predictive model attends mostly to A&E attendances (25.2%), CXR (24.3%), Breathlessness (27.1%), lung function tests, Urine examination and smoking as well as the conditions such as Diabetes (23.6%), Obesity (21.1%), other cancers, and Musculoskeletal disorder, disorder of spine, Back disorders.

The COPD cluster (11·2% of total lung cancer patients) contains over 98% of lung cancer patients with COPD and respiratory condition monitoring. 86·1% of patients are smokers, ranking the second among the six clusters. 45% of patients attended A&E and CXR. Our model attends mostly to breathlessness, CXR, A&E attendances, chest pain, COPD, Asthma, Obesity, Acute non-infective exacerbation of COPD, and other cancers. This cluster also includes patients with Cardiac and circulatory system disease.

The shorter pathway cluster (24·0% of total lung cancer patients) consists of the patients whose three-year pathways contain much fewer (10-50) medical codes than other clusters (30-398). This cluster contains the highest portion of smokers (Over 93%), 55% of patients are drinkers and 48% of patients had Chest X-ray, 21% patients received fast-track referral for suspected lung cancer. Our predictive model also attends mostly to Smoking, Chest X-ray and Fast track referral for 20%-38% patients. These figures are much more than that of the other clusters. This suggests that there may be three reasons for short diagnostic pathways: patients receiving timely referrals and efficient investigations, younger patients (age<55 yrs) who used to be healthy and seldom visited GP but developed symptoms of lung cancer or data quality issues as codes suggestive of lung cancer are less likely to be entered if cancer is not suspected. 20% to 40% of patients in this cluster presented breathlessness, nervous system symptoms, e.g. Feeling anxious, seen by A&E, suffer from COPD, Obesity, other cancers and Musculoskeletal disorder, disorder of spine, Back disorders and took blood test, lung function test, Bronchoscopy and biopsy. Our model also attends mostly to these factors for more than 10% patients.

The other chronic condition cluster only contains 2·3% of total lung cancer patients. 100% of this cluster's patients are under chronic condition monitoring (except for COPD, Respiratory, Diabetes, Hypertension, cardiac diseases). Over 80% of patients are smokers and have Cardiac disease, Circulatory system diseases. Over 60% of patients are drinkers, ranking the second among the six clusters. Our model attends mostly to Breathlessness, Lung function test, CXR, Seen by A&E, COPD, other cancers, smoking, chest pain, Obesity, Musculoskeletal disorder, disorder of spine, Back disorders, and Acute non-infective exacerbation of COPD for 8%-45% of lung cancer patients.

The Acute condition cluster (23·1% of total lung cancer patients) contains the patients who presented much more acute conditions with high proportions than the other clusters, for example, Nervous system symptoms, Feeling anxious (1st rank), Musculoskeletal disorder, disorder of spine, Back disorders, Eye conditions, Genito-Urinary system disease, Atypical fibroxanthoma of skin, Chest infection, Polymyalgia, Other skin and subcutaneous tissue disorders, Digestive system diseases, Bacteraemia, Anxiety disorder, ear condition. Our model attends mostly to Breathlessness, CXR, plain X-rays, Seen by A&E, other cancers, smoking, chest pain, Musculoskeletal disorder, disorder of spine, Back disorders for over 10% of patients.

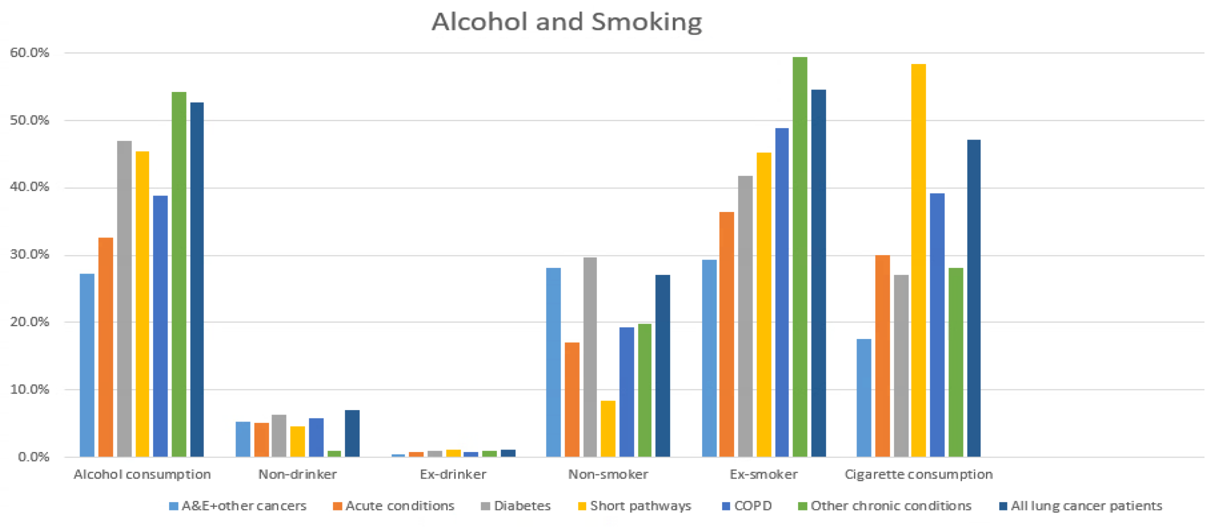


Figure S14: Alcohol and Tobacco consumption for the lung cancer patients across the six clusters.

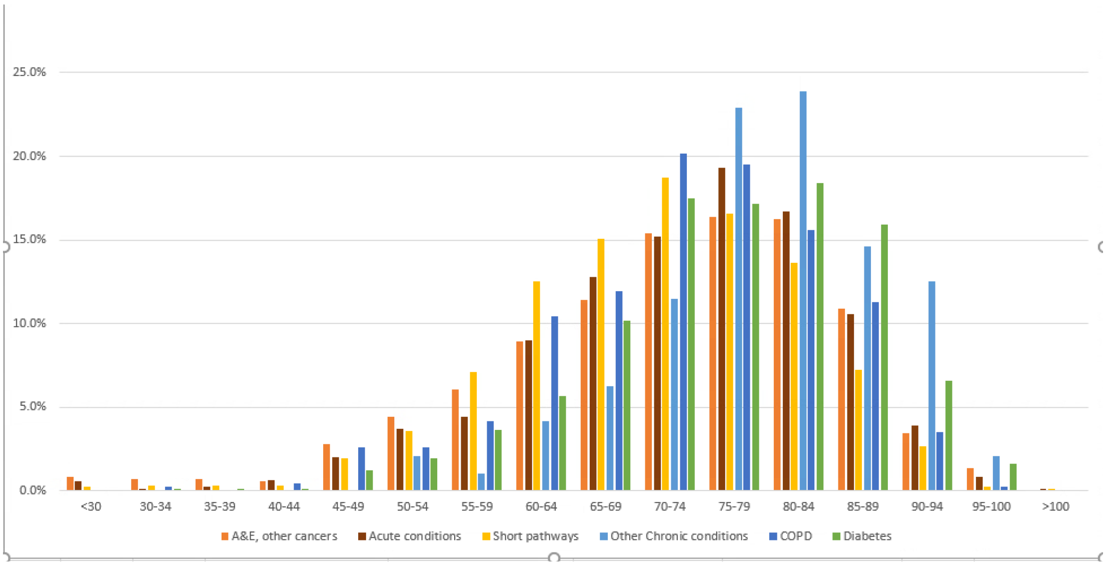

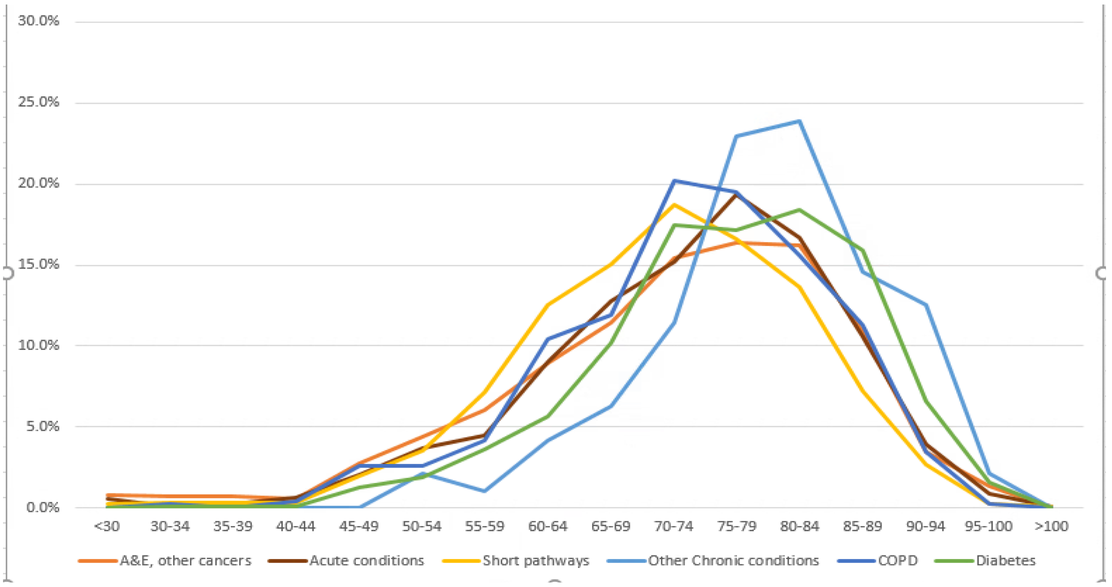


Figure S15: The distribution of lung cancer patients across different age groups in the six clusters. There are more young patients (<70 yrs) (especially patients (<55 yrs)) in the clusters of Acute conditions, Short pathways and A&E + other cancers
